# Introducing and Evaluating the Patient Report Template for AI-Powered Nursing Handoffs

**DOI:** 10.1101/2024.12.31.24319818

**Authors:** Gabriel Vald, Yusuf Sermet, Nai-Ching Chi, Ibrahim Demir

**Author notes:** Corresponding Author, Gabriel Vald.

## Abstract

This study evaluates the effectiveness of the Patient Report Template (PRT) in addressing inefficiencies in nursing workflows related to electronic health records (EHRs) and clinical decision support systems. The PRT aims to streamline patient handoffs, reduce charting time, enhance direct care hours, and improve patient safety. A survey was sent to 2,118 nurses at the University of Iowa Health Care System in order to gather feedback, with 106 participants electing to assess the perceived usefulness of the PRT components and their attitudes toward integrating artificial intelligence (AI) into clinical documentation. Participants rated sections of the PRT, including Patient Profile, Review of Systems, and Safety, on a five-point Likert scale, with most components receiving high ratings for usefulness. Comfort and trust in AI were notably low, though respondents acknowledged the potential utility of AI-generated reports. The findings highlight the PRT’s potential to reduce cognitive load, improve information consistency during handoffs, and address EHR-related challenges. Future work will involve implementing the PRT in real-world clinical settings to validate its utility & accuracy and to explore its adaptability across specialized nursing units.

**What is known:**

- Electronic health records and clinical decision support systems carry burdens associated with data retrieval and entry, as well as introduce more friction to clinical workflow.
- Electronic health record data is vast; free text clinical notes are abundant and underused.
- While crucial for care continuity, handoffs often lack standardization and thus are prone to information loss and safety risks.

**What this paper adds:**

- Creation and feedback on a clinical decision support template which aims to reduce pain points associated with charting.
- Feedback from 106 University of Iowa Health Care nurses about what they would and would not find useful in a patient handoff report.
- Pathway to further usability and accuracy testing for reports which make use of items from the patient report template.

## 1. Introduction

Research shows that nurses often spend significant time on tasks such as medication management, staff communication, and equipment preparation, which reduces direct patient care time (Schachner et al., 2015; Yen et al., 2018). Although certain non-care tasks, like organizing supplies and ongoing education, are essential, there is room for improvement in other areas (Liang et al., 2012). The use of EHRs is one area that presents both advantages and challenges for nurses (Yee et al., 2012). While EHRs allow access to critical information, like medication schedules and historical data, issues such as software unfamiliarity, system outages, and restrictive data entry methods can hinder nurses’ efficiency (Park et al., 2006).

Nurses report spending 20-50% of their day on EHRs, often limiting time for patient care (Schachner et al., 2015; Yee et al., 2012). Additionally, extended charting time has been associated with increased clinician burnout, indicating friction within data entry processes (Gardner et al., 2018). Increasing direct care time has demonstrated benefits for patient outcomes, with reduced care time linked to higher patient mortality, even when controlling for hospital and patient factors (Liang et al., 2012). While increasing staffing levels is one approach, minimizing time spent on charting via integration with clinical decision support systems could also enhance patient outcomes by freeing more time for direct care.

In the 2000s, nursing clinical decision support emerged as a distinct discipline, focusing on areas such as diagnostic support, medication management, situational awareness, guideline adherence, and non-medication interventions (e.g., memory techniques) (Lopez et al., 2016). By 2013, approximately 41% of US hospitals with EHRs also utilized clinical decision support systems (Sutton et al., 2020). However, several limitations of these systems have been identified:

*<u>Fragmented Workflows</u>*: Early systems often required documentation outside of regular workflows, increasing cognitive effort and task completion times (Sutton et al., 2020).

*<u>Alert Fatigue</u>*: Up to 95% of alerts have been found inconsequential, leading to distrust and disruptions in clinician workflows. Alerts should be reserved for critical or life-threatening situations to maintain effectiveness (Sutton et al., 2020).

*<u>Computer Literacy Dependence</u>*: Overly complex systems can hinder engagement, particularly among clinicians with limited technological proficiency. Effective training is essential to ensure confident system use (Sutton et al., 2020).

Despite these challenges, opportunities exist to develop systems that address these issues while supporting clinicians. One key gap is knowledge transfer during nursing handoffs, where lapses in communication can lead to incomplete information and patient safety risks. The lack of standardized handoff procedures contributes to inconsistent documentation and missed clinical details, affecting continuity of care (Keenan et al., 2013). Verbal communication and handwritten notes, commonly used during handoffs, are particularly prone to errors and omissions. Clinical Decision Support (CDS) systems can mitigate these issues by standardizing patient data formats, ensuring consistent information transfer, and reducing manual workload (Keenan et al., 2013). While EHRs provide access to necessary data, retrieving long-form clinical text, such as physicians’ notes, remains impractical during shift changes. Moreover, existing EHR dashboards often lack usability testing, leading to tools that fail to meet clinicians’ needs. Iterative testing and refinement are essential to address the dynamic data requirements of care teams effectively (Keenan et al., 2013).

Large language models (LLMs), such as GPT-4 and related models, are capable of handling vast amounts of text and generating coherent, concise summaries without requiring extensive fine-tuning (Tang et al., 2023; Samuel et al., 2024). This capability allows these models to sift through detailed and complex information, providing users with key insights and relevant data in a more accessible format (Sajja et al., 2023a). This is an advantage over traditional systems that struggle to extract relevant details effectively (Pursnani et al., 2023). By summarizing free-text input, LLMs enable access to critical information that might otherwise be overlooked (Van Veen et al., 2024, Sajja et al., 2023b). This capability is particularly valuable in healthcare, where extracting insights from clinical notes is essential. In nursing handoffs, where verbal communication may introduce inconsistencies, LLMs could generate summaries to cross-reference handoff details, ensuring accurate transmission of data such as lab results or treatment changes (Sermet & Demir, 2021; Van Veen et al., 2024). This added layer of support could help to maintain consistent access to essential clinical information, making LLMs a potentially powerful tool for summarizing and curating unstructured medical data.

In response to these gaps and opportunities, the Patient Report Template (PRT) has been developed as a structured tool to facilitate more effective nursing handoffs and streamline EHR documentation. By assessing nurse opinions on each component of the PRT, gaps in research related to practicality and usefulness of structured report items can be addressed. The PRT combines established handoff methods with essential patient information, creating a comprehensive and easily accessible summary designed to reduce cognitive load and improve communication among nursing staff. It is grounded in evidence-based handoff strategies such as SBAR (Situation, Background, Assessment, Recommendation) and I PASS the BATON, ensuring it aligns with existing clinical practices. Specifically, the PRT includes several key components:

*<u>Patient Profile</u>*: This section contains essential identification and demographic information, such as the patient’s name, age, sex, attending physician, and code status.

*<u>Review of Systems</u>*: A structured summary of the patient’s presenting complaints and relevant medical history, ensuring that critical clinical information is readily available.

*<u>Situation Overview</u>*: A concise description of the patient’s current clinical status and plan of care, enabling nurses to quickly assess the patient’s needs.

*<u>Safety Section</u>*: This component provides critical safety information, including allergies and lab values, to support safe patient care.

*<u>Actions and Next Steps</u>*: Documentation of interventions already performed and planned actions, enabling clear communication about the continuity of care.

This study evaluates the PRT’s potential to address inefficiencies in nursing workflows related to EHR documentation and handoffs. A survey of nurses will assess the perceived usefulness of the PRT and their perspectives on integrating AI into clinical documentation, focusing on comfort, trust, and expected utility. Feedback from a diverse group of nursing staff across departments will ensure findings reflect the perspectives of end-users. By examining the PRT’s impact, the study aims to inform the optimization of nursing practices and support patient safety through innovative tools.

## 2. Methods

To address challenges with charting time and improve accuracy of knowledge transfer between shifts, many aspects of the patient handoff that nurses complete on shift change were brought into the PRT. The template presented in this paper represents a combination of common handoff methods (such as SBAR, I PASS the BATON; Riesenberg et al., 2010), with additional supplemental information like review of systems which is standard for both nurses and physicians. By combining patient handoff information, review of systems, and the patient plan of care, the PRT aims to provide an at a glance, factual summary for nurses, where exact values can be trusted as accurate, and the most up-to-date information is readily available at the time of care. Given that nurses see many patients per shift, the mental load of remembering the handoff for all of them can be taxing, and may lead to additional trips to the EHR in order to verify information. By directly addressing current problems in nursing, the PRT aims to be integrated into existing workflows, where a nurse can glance at a report before visiting a patient, replacing the nurse browsing the EHR for the same information.

### 2.1 Patient Report Template

The following section outlines the methods for the development of the PRT, each component’s basis and content are outlined.

#### 2.1.1 Patient Profile

The first piece of the PRT is Patient Profile. This is all relevant information pertaining to a patient’s identification. The patient’s Name, Age, Sex, Attending Physician, Identifiers (such as Medical Record Number (MRN), date of birth, or other unique identifiers), Location in the medical facility (room number, unit, bed), Code Status (e.g., Do Not Resuscitate), Isolation Status (e.g., Contact Isolation, Droplet Isolation). These pieces of information together serve to provide all of the mostly used identification information for a particular patient. An example of a Patient Profile is shown in Table 1.

**Table 1.** Patient Profile Section and Example Data.

| Component | Example Data |
| --- | --- |
| Name | *Jane Doe* |
| Age | *60* |
| Sex | *Female* |
| Physician | *Dr. John Smith, M.D.* |
| Identifiers | *MRN: 0000-00-0000*<br>*DOB: 01/01/1964* |
| Location | *Neurology Unit, Room 123* |
| Code Status | *DNR* |
| Isolation Status | *None* |

#### 2.1.2 ROS/Chief Complaint

The Chief Complaint & Review of Systems (ROS) section in the PRT is designed to align with existing clinical communication practices, facilitating its meaningful use by both nursing and medicine. This section includes the Chief Complaint (primary issue or symptom) and the following ROS items: Constitutional (general health, fever, fatigue), Eyes (ocular diseases, vision changes), ENT (ear, nose, and throat conditions), Cardiovascular (heart diseases, chest pain), Respiratory (cough, shortness of breath), Gastrointestinal (GI disorders, abdominal pain), Genitourinary (UTIs, renal conditions), Musculoskeletal (joint pain, muscle weakness), Integumentary (dermatologic conditions, rashes), Neurological (stroke, epilepsy, headaches), Psychiatric (mental health conditions, anxiety, sleep issues), Endocrine (diabetes, thyroid disorders), Hematologic / Lymphatic (blood disorders, lymphadenopathy), and Allergic / Immunologic (allergies, autoimmune diseases). Placed early in the PRT, this section enables nurses to quickly access critical patient information without searching or scrolling. By using terminology familiar to both nursing and medicine and adhering to guidelines from the American College of Cardiology (American College of Cardiology, n.d.), the PRT integrates seamlessly into established workflows. Table 2 provides an example of the Review of Systems section.

**Table 2.**
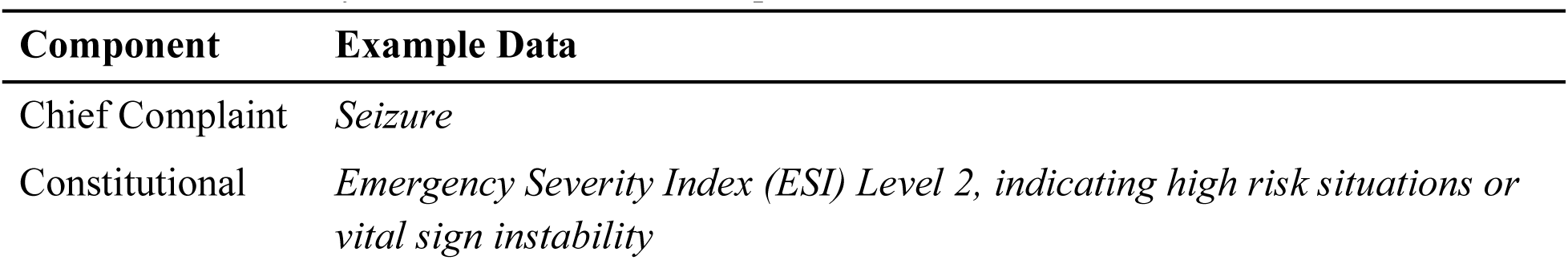
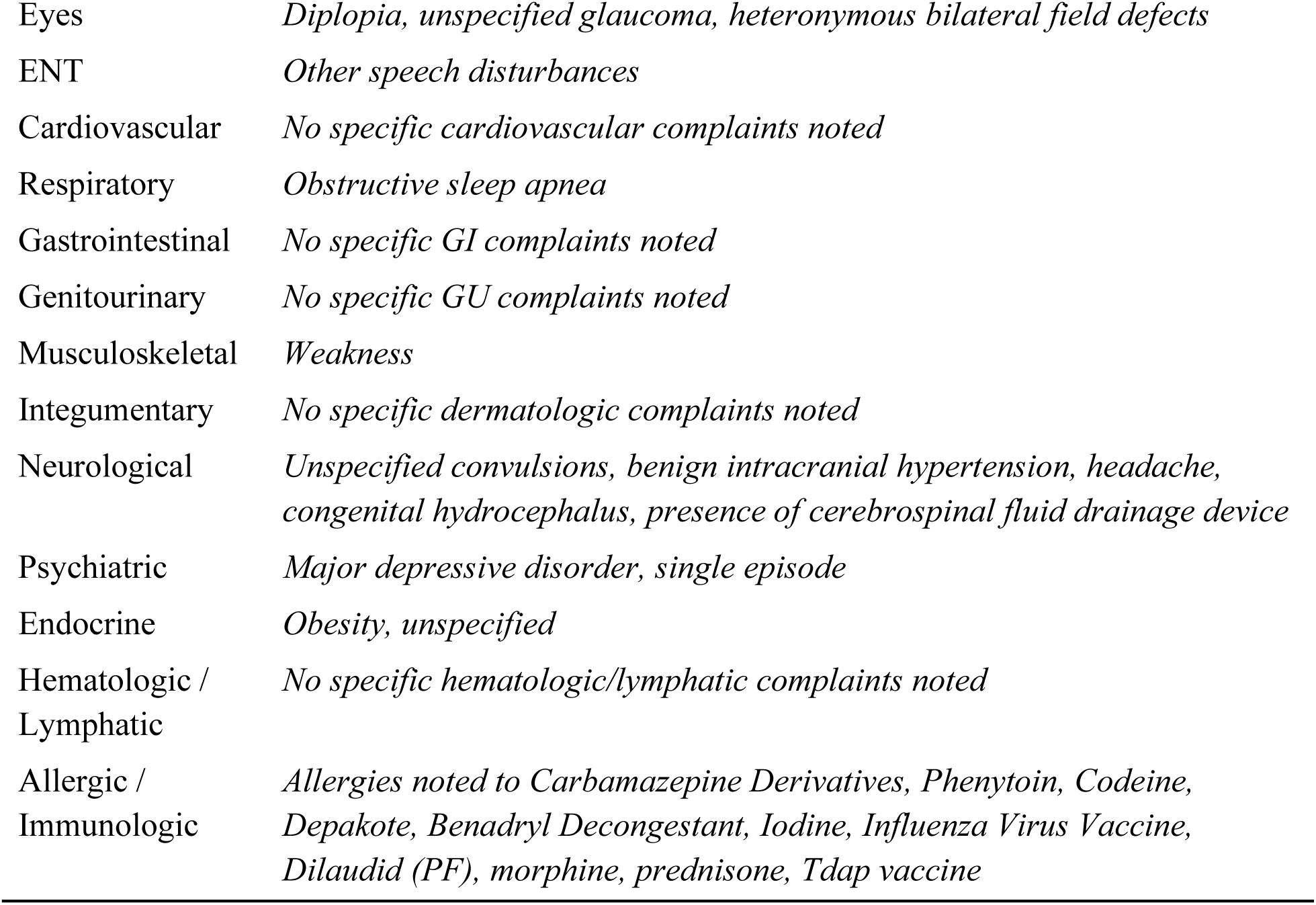
Review of Systems Section and Example Data.

| Component | Example Data |
| --- | --- |
| Chief Complaint | *Seizure* |
| Constitutional | *Emergency Severity Index (ESI) Level 2, indicating high risk situations or vital sign instability* |
| Eyes | *Diplopia, unspecified glaucoma, heteronymous bilateral field defects* |
| ENT | *Other speech disturbances* |
| Cardiovascular | *No specific cardiovascular complaints noted* |
| Respiratory | *Obstructive sleep apnea* |
| Gastrointestinal | *No specific GI complaints noted* |
| Genitourinary | *No specific GU complaints noted* |
| Musculoskeletal | *Weakness* |
| Integumentary | *No specific dermatologic complaints noted* |
| Neurological | *Unspecified convulsions, benign intracranial hypertension, headache, congenital hydrocephalus, presence of cerebrospinal fluid drainage device* |
| Psychiatric | *Major depressive disorder, single episode* |
| Endocrine | *Obesity, unspecified* |
| Hematologic /<br>Lymphatic | *No specific hematologic/lymphatic complaints noted* |
| Allergic /<br>Immunologic | *Allergies noted to Carbamazepine Derivatives, Phenytoin, Codeine, Depakote, Benadryl Decongestant, Iodine, Influenza Virus Vaccine, Dilaudid (PF), morphine, prednisone, Tdap vaccine* |

#### 2.1.3 Situation

The Situation section of the PRT derives its name from the "S" in the SBAR handoff method (Shahid & Thomas, 2018) and is also included in the I PASS the BATON method (Riesenberg et al., 2010). This section provides a quick summary of the current clinical situation and includes four components: Level of Uncertainty (quantifying uncertainty, possible differential diagnoses), Recent Changes (updates to the patient’s condition), Response to Treatments (patient’s reaction to interventions), and Plan of Care (current nursing care plan). Positioned near the beginning of the PRT, following the Chief Complaint and ROS sections, it prioritizes the patient’s current state to facilitate effective handoffs. Table 3 presents an example of the Situation section.

**Table 3.**
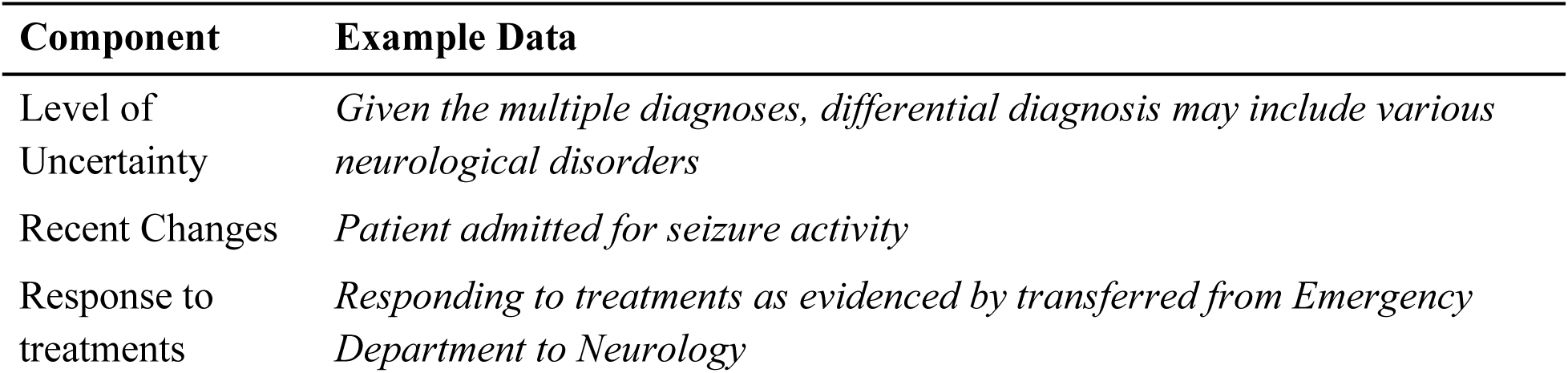
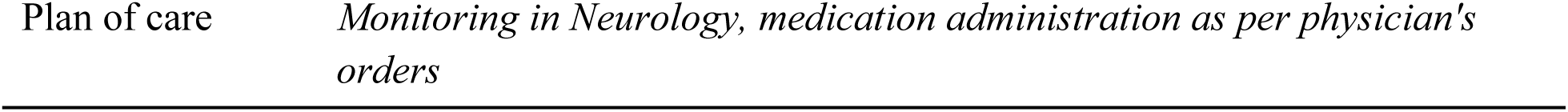
Situation Section and Example Data.

| **Component** | **Example Data** |
| --- | --- |
| Level of<br>Uncertainty | *Given the multiple diagnoses, differential diagnosis may include various neurological disorders* |
| Recent Changes | *Patient admitted for seizure activity* |
| Response to<br>treatments | *Responding to treatments as evidenced by transferred from Emergency Department to Neurology* |
| Plan of care | *Monitoring in Neurology, medication administration as per physician's orders* |

#### 2.1.4 Safety

While not explicitly stated earlier, the PRT also aims to reduce errors and safety events in addition to cognitive load reduction and clinical decision support. The Safety section addresses this goal by presenting critical patient information. Lab Values are reported first, with anomalous values prioritized and routine values listed separately in reverse chronological order. Allergies are listed next, covering medication, food, or other relevant allergies. Lastly, Alerts such as fall risks, isolation requirements, or other critical notifications are included. This section serves as a convenient cross-reference for clinicians, eliminating the need to manually retrieve this information from the EHR during care. Table 4 outlines the Safety section’s structure.

**Table 4.** Safety Section and Example Data.

| Component | Example Data |
| --- | --- |
| **Lab Values** | *Bicarbonate: 21.0 (low: 22.0, high: 32.0) [abnormal]*<br>*MCHC: 30.2 (low: 32.0, high: 37.0) [abnormal]*<br>*Glucose: 128.0 (low: 70.0, high: 100.0) [abnormal]*<br>*RDW-SD: 50.8 (low: 35.1, high: 46.3) [abnormal]* |
| **Allergies** | *Carbamazepine Derivatives, Phenytoin, Codeine, Depakote, Benadryl*<br>*Decongestant, Iodine, Influenza Virus Vaccine, Dilaudid (PF), morphine,*<br>*prednisone, Tdap vaccine* |
| **Alerts** | *None specified* |

#### 2.1.5 Background

The Background section also comes originally from the SBAR & I PASS the BATON format of nursing handoffs and aims to give all relevant background information related to a patient’s care (Riesenberg et al., 2010). The items in the Background section are Comorbidities (other diseases which are simultaneously present in the patient besides the Chief Complaint), Previous Episodes (previous hospitalizations, surgeries, illnesses, complications), Current Medications (current list of medications used by patient and relevant medications ceased), and Family History (e.g., Hereditary conditions, history of chronic disease, addiction risk). These individual components, when combined, provide a detailed picture of a patient’s history and any potential inherited risk factors. Table 5 shows an example Background section.

**Table 5.** Background Section and Example Data.

| Component | Example Data |
| --- | --- |
| **Comorbidities** | *Asthma, chronic stable*<br>*Pruritus*<br>*Pseudotumor cerebri*<br>*Chronic headache*<br>*Seizures vs pseudoseizures* |
| **Previous Episodes** | *Previous hospitalizations for congenital hydrocephalus, vertigo, dizziness, and pseudoseizures*<br>*Previous surgeries including cochlear implants, VP shunt, LS shunt, umbilical hernia repair, laparoscopic cholecystectomy* |
| **Current Medications** | *Acetaminophen, azelastine, albuterol sulfate, montelukast, fluticasone, omeprazole, ibuprofen, acetazolamide, cholestyramine, topiramate, lorazepam, sodium chloride flush* |
| **Family History** | *Neurology note reports seizures in mother after head trauma* |

#### 2.1.6 Actions

The actions section of the PRT aims to provide an accounting of previous & upcoming actions to be taken related to a patient, the rationale, and when to fetch the physician upon changes in patient condition. The sections for action include Actions Taken (Detail the actions and interventions that have already been completed), Actions Required/Ongoing (Explain any treatments or procedures currently in progress or that still need to be completed), Rationale (justification for previous and current/future actions), and finally Signs to Elevate (symptoms which indicate need to alert physician/nurse practitioner). This section aims to both allow nurses to see what has and needs to be done, but also aims to give them a sense of when they should call the doctor in case of a specific relevant symptom or change in condition. Table 6 contains an example of the Actions section.

**Table 6.**
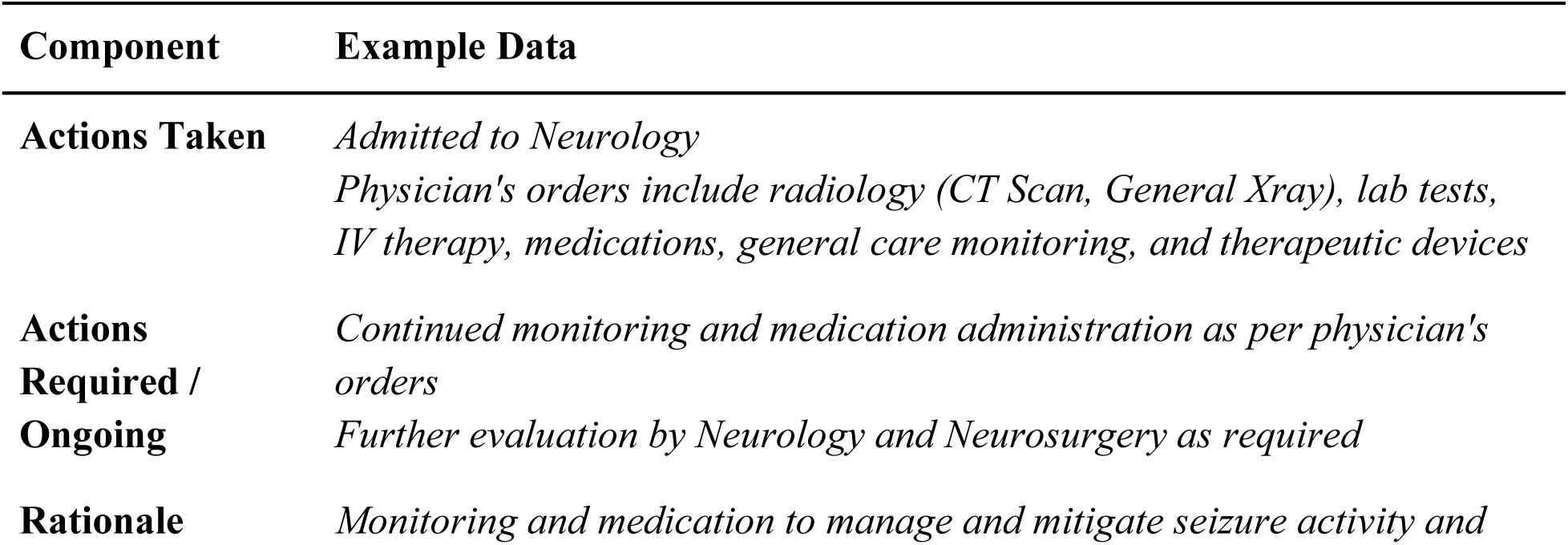
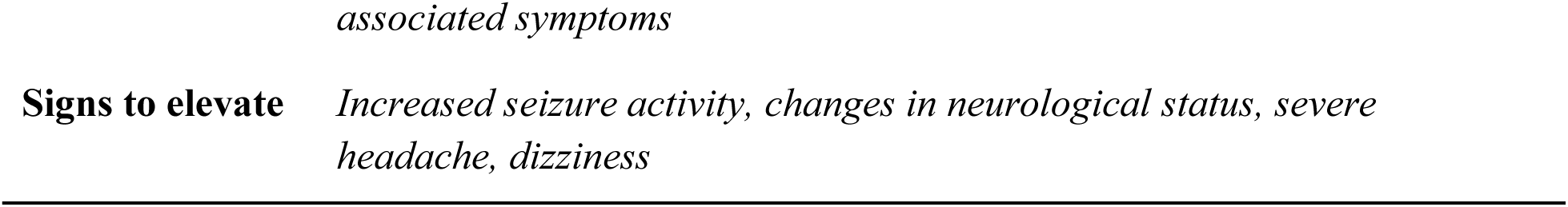
Actions Section and Example Data.

#### 2.1.7 Timing

The timing section aims to give nurses a guide for caring for the patient. It contains both explicit timing and ordinal instructions, which specify the order in which tasks should be completed. The Timing section is comprised of Levels of Urgency/Prioritization (order of actions of care in decreasing priority), Explicit Timing (if specific times are required for certain actions, e.g., administer medicine, remove ventilator), and Coordination (mention any coordination with other departments or services, e.g., such as x-ray, lab work). Example data for the Timing section are contained in Table 7.

**Table 7.**
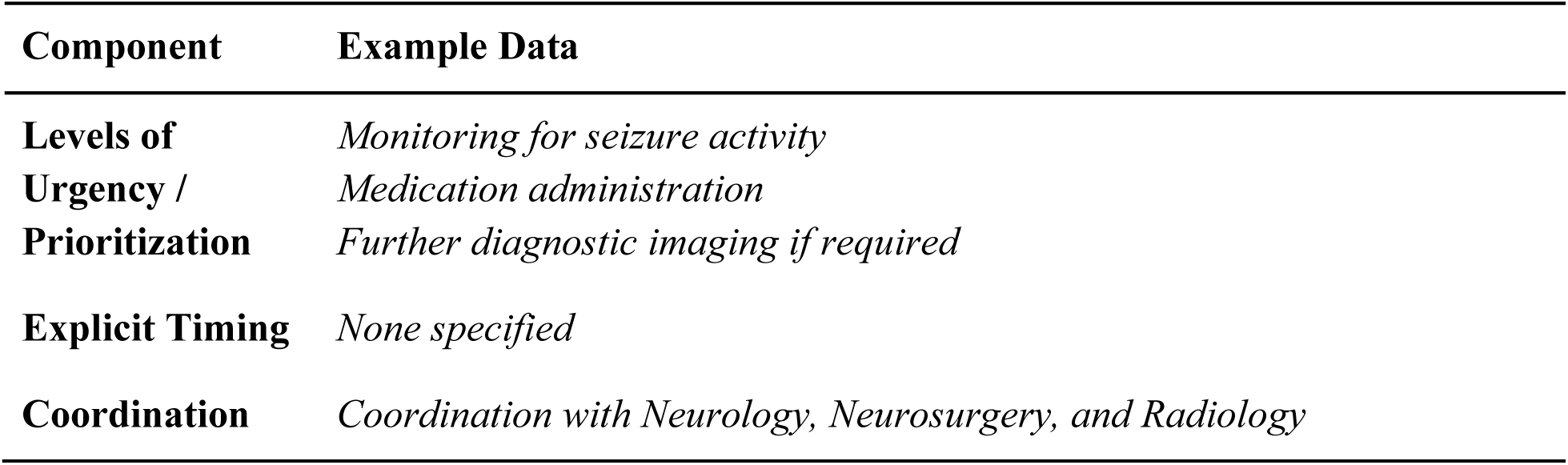
Timing Section and Example Data.

| Component | Example Data |
| --- | --- |
| **Levels of Urgency / Prioritization** | *Monitoring for seizure activity*<br>*Medication administration*<br>*Further diagnostic imaging if required* |
| **Explicit Timing** | *None specified* |
| **Coordination** | *Coordination with Neurology, Neurosurgery, and Radiology* |

#### 2.1.8 Ownership

The ownership section was designed to give nurses a clear idea about those who are most related to the patient’s care. The sections include Responsibility (What person/team is in charge of the patient’s care) and Patient/Family (Who is making medical decisions for the patient). By clearly outlining these two details, nurses are able to view relevant contacts for the patient both in terms of managing care, and in terms of communicating with relevant family members or the patient themself regarding their status or care. Table 8 contains an example of the Ownership section.

**Table 8.** Ownership Section and Example Data.

| Component | Example Data |
| --- | --- |
| **Responsibility** | *Neurology team in charge of care* |
| **Patient/Family** | *Medical decisions to be made by patient or designated family member* |

#### 2.1.9 Next

The next section addresses the expected course for the patient into the future. This includes the Anticipated Changes (Any possible/likely changes in status for the patient as a result of the interventions/care), the Plan of Action (plan of care for the patient if current conditions persist), and any Contingency Plans (If current conditions change in specific ways, adaptations to plan or other plans of care for the patient). The information that can be expected in the Next section is contained in table 9.

**Table 9.** Next Section and Example Data.

| Component | Example Data |
| --- | --- |
| **Anticipated Changes** | *Possible changes in neurological status depending on response to interventions* |
| **Plan of action** | *Continued monitoring and medication administration* |
| **Contingency Plans** | *If seizure activity increases, escalate care to more intensive monitoring or intervention*<br>*If neurological status changes, re-evaluate treatment plan* |

### 2.2 Survey of Nurses

Before developing a comprehensive system that combines LLMs and EHR, it’s important to understand whether or not nurses would find utility in such a system. In order to assess the demand for the PRT as well as understand nurses’ sentiments towards artificial intelligence (Zhang et al., 2023), its expected value in their work, and their trust in such systems. A survey was chosen as the data collection method for this study, as it facilitated the largest possible body of feedback. The cost-effectiveness of a survey further enabled a broad reach. The use of a survey also helps to guarantee anonymity, which enables participants to offer their opinion more freely, without the fear of disappointing or upsetting researchers. Standardized, efficient data collection are also chief reasons for pursuing a survey for this study, ensuring all nurses answer the same questions on the same scales.

#### 2.2.1 Participants

A mass-mail email system was used to invite currently practicing nurses from the University of Iowa Health Care (UIHC) system to participate in the survey. UIHC is an academic medical center providing general acute care and specialized services. Its facilities include a main hospital, the UI Stead Family Children’s Hospital with 205 pediatric beds, the UI Health Care Downtown, and regional outreach clinics across Iowa. Annually, UIHC handles over 1.2 million outpatient visits, 33,000 inpatient admissions, and 52,000 emergency department visits, with more than 36,000 major and 190,000 minor surgical procedures (UIHC, n.d.). Its nursing staff, totaling over 5,300, has earned five Magnet designations for excellence in patient care. UIHC integrates clinical care, research, and education to serve Iowa and the surrounding region. No departmental requirements were included in the mass-mail cohort, so nurses from all units received the recruitment offer.

#### 2.2.2 Survey Design

To evaluate nurses’ perceptions of AI in their clinical workflows and assess the utility of the PRT, a survey was developed with two objectives: (1) to understand nurses’ comfort, trust, and perceived utility of AI-generated reports, and (2) to determine the usefulness of the PRT for patient handoffs and clinical documentation. Nurses rated their comfort, trust, and expected utility of AI on a five-point Likert scale, ranging from "Not at all" to "Extremely" or "No utility" to "Exceptional utility." They also evaluated the PRT’s components, including the patient profile, review of systems, situation overview, safety alerts, and action plans, on a similar scale to gauge its relevance in daily workflows. The survey also collected demographic data, including department, years of experience (in 10-year increments), and age group (Young Adult, Adult, Older Adult). It was reviewed by a practicing nurse for clarity and revised accordingly. Distributed via email to 2,118 nurses at the University of Iowa Healthcare system, participation was voluntary and anonymized. A follow-up email and a $5 incentive ensured broader participation while maintaining privacy through separate payment forms unlinked to survey responses. This study was approved by the University of Iowa Institutional Review Board (IRB # 202407425) with Exempt status.

## 3. Results

Below is the report on the results of the survey. The following section covers the PRT, Nurses’ sentiments towards AI in their work, and the results of the 3 demographic questions that the nurses were surveyed on.

### 3.1 Patient Report Template

The following section contains the results of Nurses’ ratings for each component of the PRT. Descriptive statistics and response distributions are given for every component.

#### 3.1.1 Patient Profile

The first section of the PRT that nurses were asked to rate for its usefulness was the Patient Profile section. The results of the Nurse’s ratings are displayed in Table 10.

**Table 10.** Nurses’ ratings of the Patient Profile section.

| Patient Profile | Mode | Median | Average | Minimum | Maximum | Count |
| --- | --- | --- | --- | --- | --- | --- |
| Name: Patient's Name | 5 | 5 | 4.42 | 1 | 5 | 106 |
| Age: Patient's age | 5 | 5 | 4.25 | 1 | 5 | 106 |
| Sex: Biological Sex of the patient | 5 | 4 | 4.03 | 1 | 5 | 105 |
| Physician: Attending physician | 5 | 4 | 4.16 | 1 | 5 | 106 |
| Identifiers: e.g., Medical Record Number (MRN), date of birth (DOB) | 5 | 5 | 4.34 | 1 | 5 | 105 |
| Location: Location within the healthcare facility (e.g., room number, unit) | 5 | 4 | 4.20 | 1 | 5 | 106 |
| Code Status: e.g., DNR | 5 | 5 | 4.38 | 1 | 5 | 106 |
| Isolation Status: e.g., Contact isolation, droplet isolation | 5 | 5 | 4.28 | 1 | 5 | 106 |

Overall, nurses found the components of the Patient Profile to be useful, with all scores achieving an average score above 4 (Very Useful) on the 5-point Likert scale. The middle (median) scores for each section range between 4 and 5 (Very & Extremely Useful), and the mode for all sections is 5 (Extremely useful). Patient’s Name and Code Status scored the highest averages, at 4.42 and 4.38 respectively. The section that scored the lowest in the Patient Profile was Sex, averaged at 4.03. Figure 1 is the distribution of answers for the Patient Profile.

**Figure 1.**
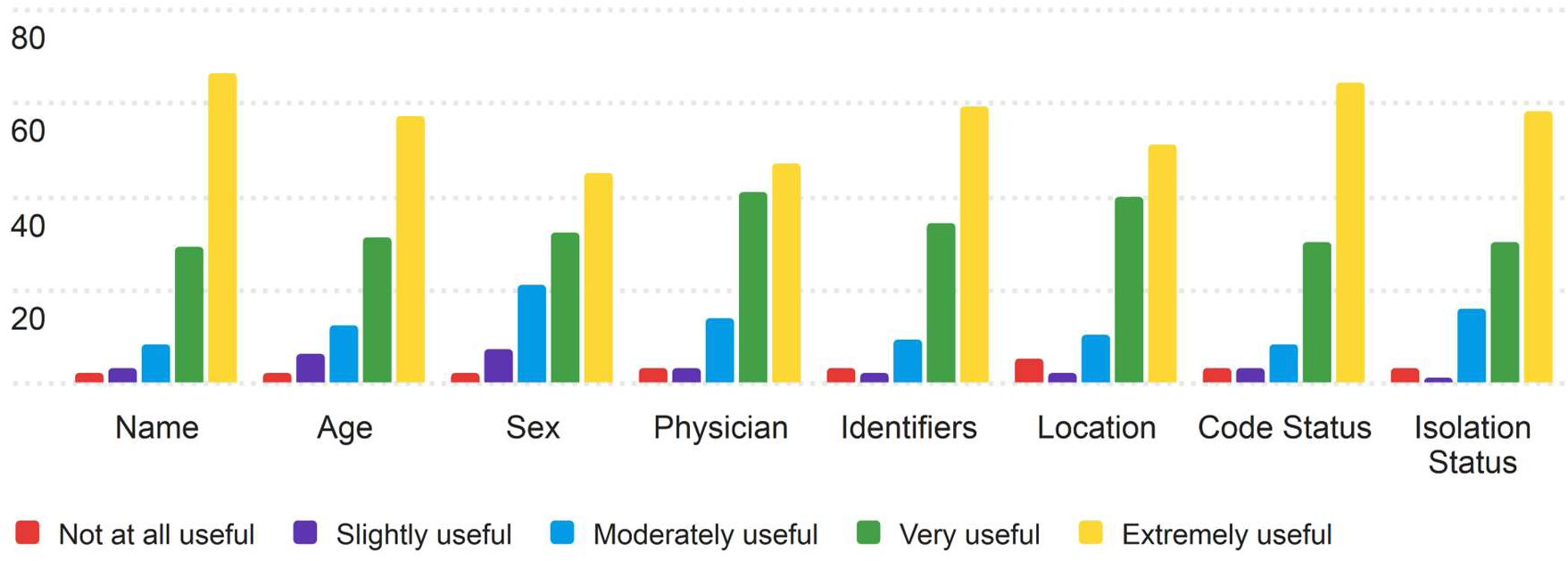
Response Distribution for the Usefulness of the Patient Profile section.

Most nurses thought that the Patient Profile section of the PRT would be useful to them in their work. In all cases, extremely useful (5 on the Likert scale) was the most common rating, with moderately useful and very useful (3 and 4 on the Likert scale, respectively) following as the next most common ratings. Each section received either 106 or 105 total ratings for the patient profile section.

#### 3.1.2 Review of Systems

The second section of the PRT that nurses were asked to rate for its usefulness was the Review of Systems section. The results of the Nurse’s ratings are displayed in Table 11.

**Table 11.** Nurses’ ratings of the Review of Systems section.

| **Review of Systems** | **Mode** | **Median** | **Average** | **Minimum** | **Maximum** | **Count** |
| --- | --- | --- | --- | --- | --- | --- |
| Chief Complaint: The primary issue or symptoms that brought the patient into care or the main concern for the current admission | 5 | 5 | 4.4 | 1 | 5 | 106 |
| Integumentary: Dermatologic conditions, breast cancer, Rashes, skin changes, breast lumps | 5 | 4 | 3.88 | 1 | 5 | 106 |
| Neurological: Neurological disorders (e.g., stroke, epilepsy), Headaches, seizures, numbness | 5 | 4 | 4.14 | 1 | 5 | 106 |
| Psychiatric: Mental health conditions, Depression, anxiety, sleep issues | 5 | 4 | 3.98 | 1 | 5 | 106 |
| Endocrine: Endocrine disorders (e.g., diabetes, thyroid), Polyuria, weight changes, intolerance | 5 | 4 | 3.99 | 1 | 5 | 106 |
| Hematologic/Lymphatic: Blood disorders (e.g., anemia, clotting issues), Bruising, bleeding, lymphadenopathy | 5 | 4 | 4.08 | 1 | 5 | 106 |
| Allergic/Immunologic: Allergies, immunodeficiencies, autoimmune diseases, Allergies, recurrent infections | 5 | 5 | 4.18 | 1 | 5 | 106 |
| Constitutional: General health overview, systemic issues, Fever, weight loss, fatigue, vitals | 5 | 5 | 4.12 | 1 | 5 | 106 |
| Eyes: Ocular diseases, systemic conditions, Vision changes, pain, redness | 5 | 4 | 3.73 | 1 | 5 | 106 |
| ENT: ENT conditions, infections, allergies, Hearing loss, congestion, sore throat | 5 | 4 | 3.85 | 1 | 5 | 106 |
| Cardiovascular: Heart diseases (e.g., MI, heart failure), Chest pain, palpitations, dyspnea | 5 | 5 | 4.23 | 1 | 5 | 106 |
| Respiratory: Respiratory conditions (e.g., asthma, pneumonia), Cough, shortness of breath, wheezing | 5 | 5 | 4.24 | 1 | 5 | 106 |
| Gastrointestinal: GI disorders (e.g., infections, IBD), Nausea, vomiting, abdominal pain | 5 | 4 | 3.93 | 1 | 5 | 106 |
| Genitourinary: UTIs, renal conditions, reproductive health, Dysuria, hematuria, pelvic pain | 5 | 4 | 3.89 | 1 | 5 | 106 |
| Musculoskeletal: Musculoskeletal disorders (e.g., arthritis), Joint pain, muscle weakness | 5 | 4 | 3.87 | 1 | 5 | 106 |

Nurses found the Review of Systems section of the PRT to be useful, though some categories scored higher than others. The median scores ranged between 4 and 5 (Very Useful to Extremely Useful), with all categories having a mode of 5, indicating that the most common rating was "Extremely Useful." The Chief Complaint, Cardiovascular, and Respiratory components received some of the highest ratings, with averages of 4.40, 4.23, and 4.24, respectively. Conversely, the Eyes section received the lowest average score, at 3.73, followed by Integumentary and Musculoskeletal, which also scored below 4 on average. These lower-scoring sections may indicate that some nurses did not find these categories critical or useful for their daily workflows. Figure 2 contains the distributions of responses for the review of the systems section.

**Figure 2.**
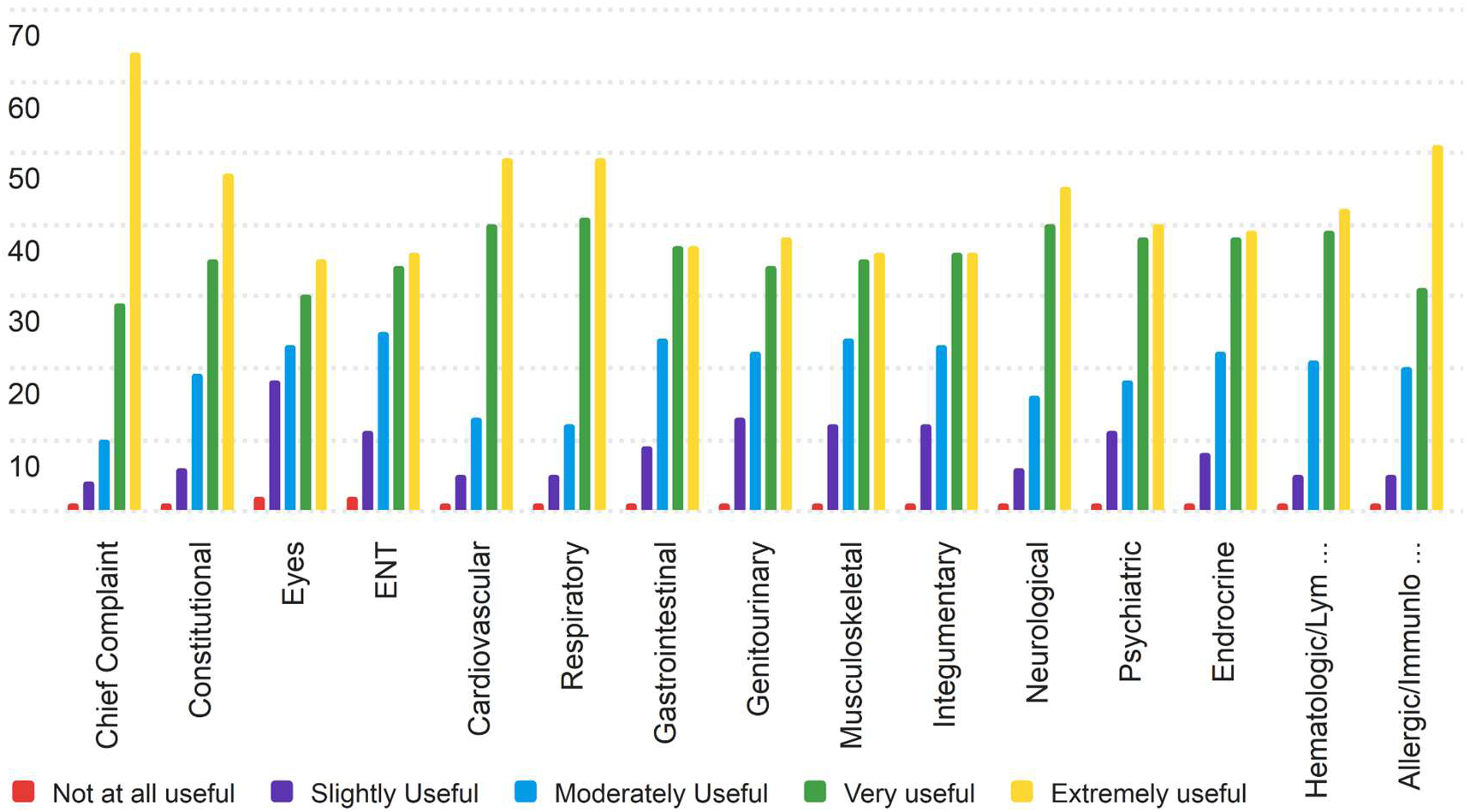
Response Distribution for the Usefulness of the Review of Systems section.

The Chief Complaint section received the highest average score (4.40), reflecting its perceived importance in nurses’ daily workflows. The lower-rated sections, such as Eyes and Integumentary, show a broader spread of responses, with fewer nurses rating them as "Extremely Useful," aligning with their lower average scores as seen in the table above.

#### 3.1.3 Situation

The third section of the PRT that nurses were asked to rate for its usefulness was the situation section. The results of the Nurse’s ratings are displayed in Table 12.

**Table 12.**
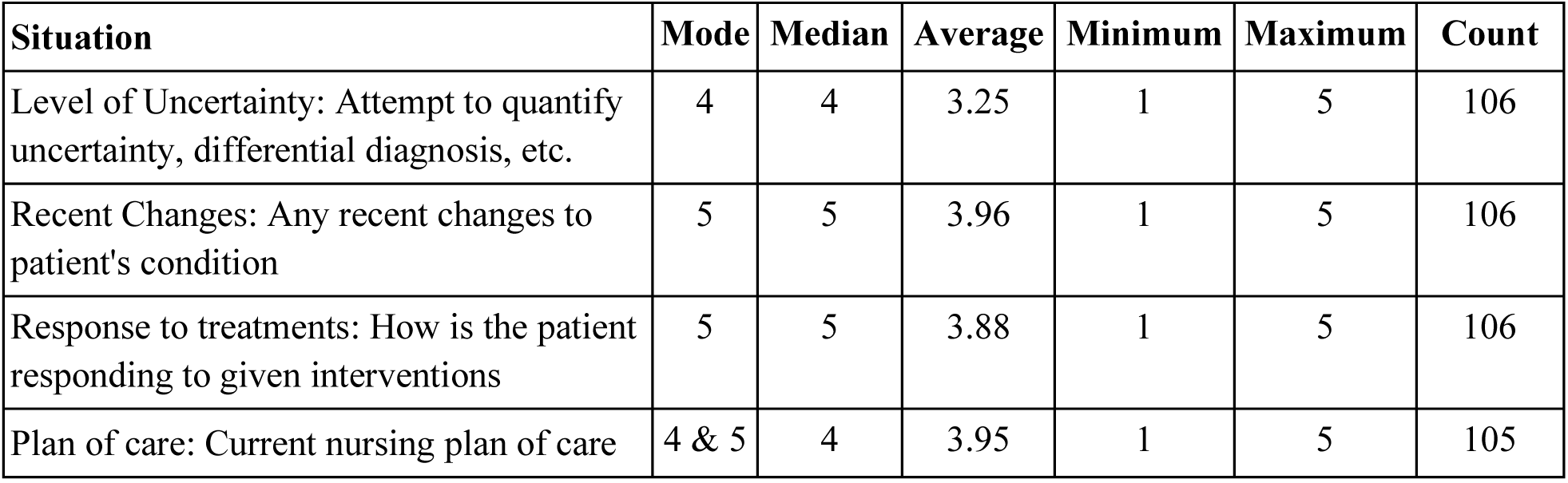
Nurses’ ratings of the Situation Section.

| Situation | Mode | Median | Average | Minimum | Maximum | Count |
| --- | --- | --- | --- | --- | --- | --- |
| Level of Uncertainty: Attempt to quantify uncertainty, differential diagnosis, etc. | 4 | 4 | 3.25 | 1 | 5 | 106 |
| Recent Changes: Any recent changes to patient's condition | 5 | 5 | 3.96 | 1 | 5 | 106 |
| Response to treatments: How is the patient responding to given interventions | 5 | 5 | 3.88 | 1 | 5 | 106 |
| Plan of care: Current nursing plan of care | 4 & 5 | 4 | 3.95 | 1 | 5 | 105 |

This sample of nurses found the Situation section to be moderately useful, with average scores ranging from 3.25 to 3.96. The Recent Changes and Plan of Care categories received the highest ratings, both with a median of 5 (Extremely Useful), suggesting these elements are highly relevant to nurses’ workflows. Level of Uncertainty had the lowest average score (3.25), indicating that this component was viewed as less useful compared to others.

The distribution of responses shows that "Very Useful" and "Extremely Useful" were common ratings for Recent Changes and Response to Treatments, while Level of Uncertainty saw more varied responses, including some lower scores. This variation in the Level of Uncertainty reflects the broader range of opinions on its relevance in daily practice. Figure 3 shows the distributions of responses.

**Figure 3.**
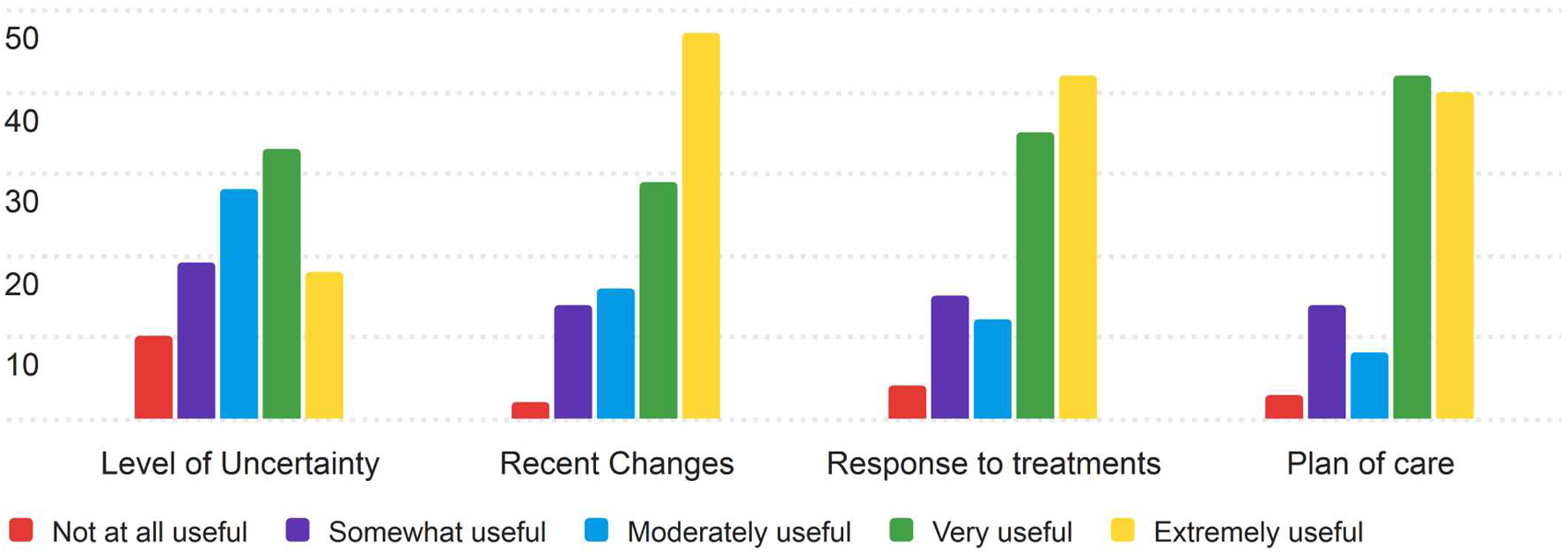
Response Distribution for the Usefulness of the Situation Section.

The Level of Uncertainty portion of the Situation section was the most controversial, with 10 nurses rating the section Not at all useful (1 on the Likert scale). Nurses deemed these sections to be less useful than prior sections, although all measures of average still see nurses rating these sections as between moderately useful and extremely useful (3-5 on the Likert scale).

#### 3.1.4 Safety

The fourth section of the PRT that nurses were asked to rate for its usefulness was the Safety section. The results of the nurses’ ratings are displayed in Table 13.

**Table 13.** Nurses’ ratings of the Safety section.

| Safety | Mode | Median | Average | Minimum | Maximum | Count |
| --- | --- | --- | --- | --- | --- | --- |
| Lab Values: Results of labs for the patient, anomalous labs first, routine results separate, chronological from most recent | 5 | 4 | 4.21 | 1 | 5 | 106 |
| Allergies: Any medication, food, or other allergies relevant to care | 5 | 5 | 4.36 | 1 | 5 | 106 |
| Alerts: fall, isolation, etc. | 5 | 4 | 4.22 | 1 | 5 | 106 |

The Safety section was found to be useful, with all components achieving an average score above 4 (Very Useful) on the 5-point Likert scale. Allergies received the highest average score of 4.36, with a median and mode of 5 (Extremely Useful), indicating strong agreement about its importance in patient safety. Lab Values and Alerts also received strong scores, with averages of 4.21 and 4.22, respectively. Most nurses rated the Safety section elements as "Extremely Useful," with the majority of remaining responses in the "Very Useful" or "Moderately Useful" categories. The distribution of responses reflects this, as seen in Figure 4.

**Figure 4.**
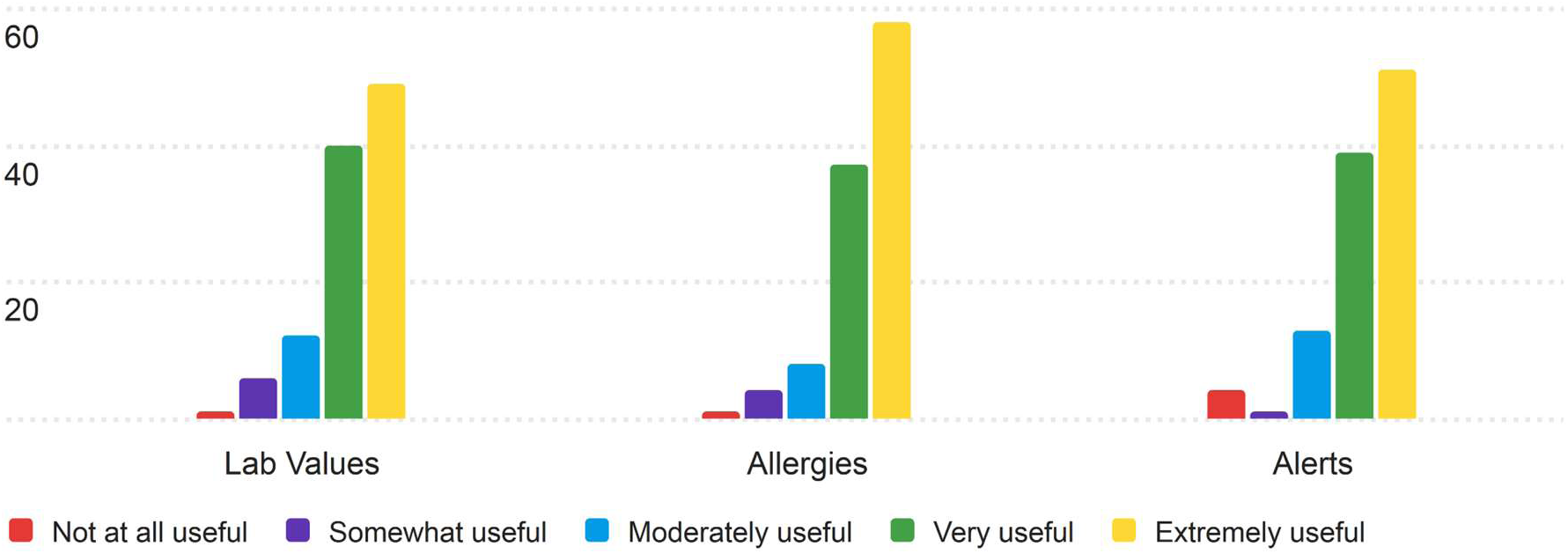
Response Distribution for the Usefulness of the Safety section.

Most nurses believe that the Safety section would be useful to their day to day work, with very few nurses rating the individual components as not at all or only somewhat useful (1 and 2 on the Likert scale), indicating that components in this section are believed to be useful across nursing disciplines.

#### 3.1.5 Background

The fifth section of the PRT that nurses were asked to rate for its usefulness was the Background section. The results of the nurses’ ratings are displayed in Table 14.

**Table 14.**
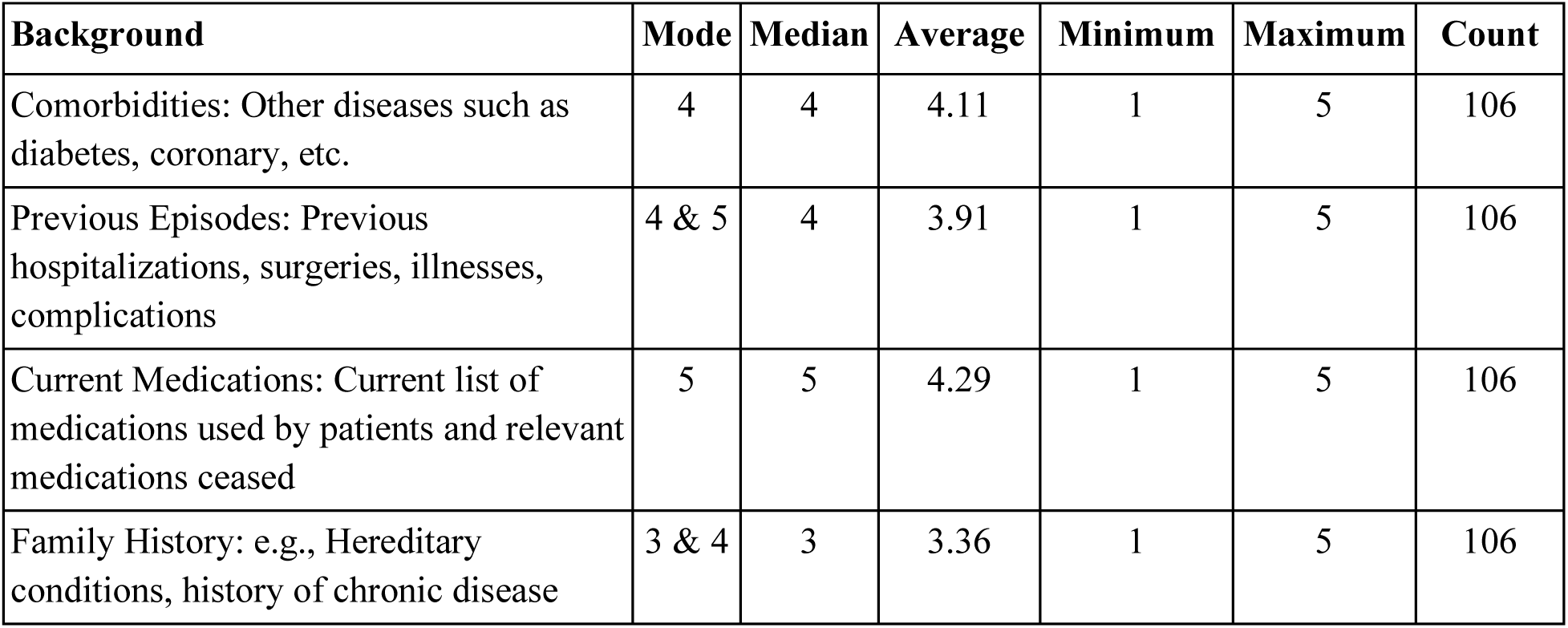
Nurses’ ratings of the Background Section.

| Background | Mode | Median | Average | Minimum | Maximum | Count |
| --- | --- | --- | --- | --- | --- | --- |
| Comorbidities: Other diseases such as diabetes, coronary, etc. | 4 | 4 | 4.11 | 1 | 5 | 106 |
| Previous Episodes: Previous hospitalizations, surgeries, illnesses, complications | 4 & 5 | 4 | 3.91 | 1 | 5 | 106 |
| Current Medications: Current list of medications used by patients and relevant medications ceased | 5 | 5 | 4.29 | 1 | 5 | 106 |
| Family History: e.g., Hereditary conditions, history of chronic disease | 3 & 4 | 3 | 3.36 | 1 | 5 | 106 |

Overall, nurses found the Background section to be moderately useful, though there was more variation in the ratings compared to other sections. Current Medications received the highest average score of 4.29, with a mode of 5 (Extremely Useful) and a median of 5, reflecting its importance in clinical practice. In contrast, Family History had the lowest average score of 3.36, with a mode of 3 and 4, indicating that nurses found this section less useful for patient handoffs.

Most ratings for Comorbidities and Previous Episodes clustered around 4 (Very Useful), with fewer responses in the "Extremely Useful" range. The Family History section had the most diverse distribution of ratings, with several responses falling in the "Somewhat Useful" and "Moderately Useful" categories. Displayed in Figure 5 are the response distributions for the background section.

**Figure 5.**
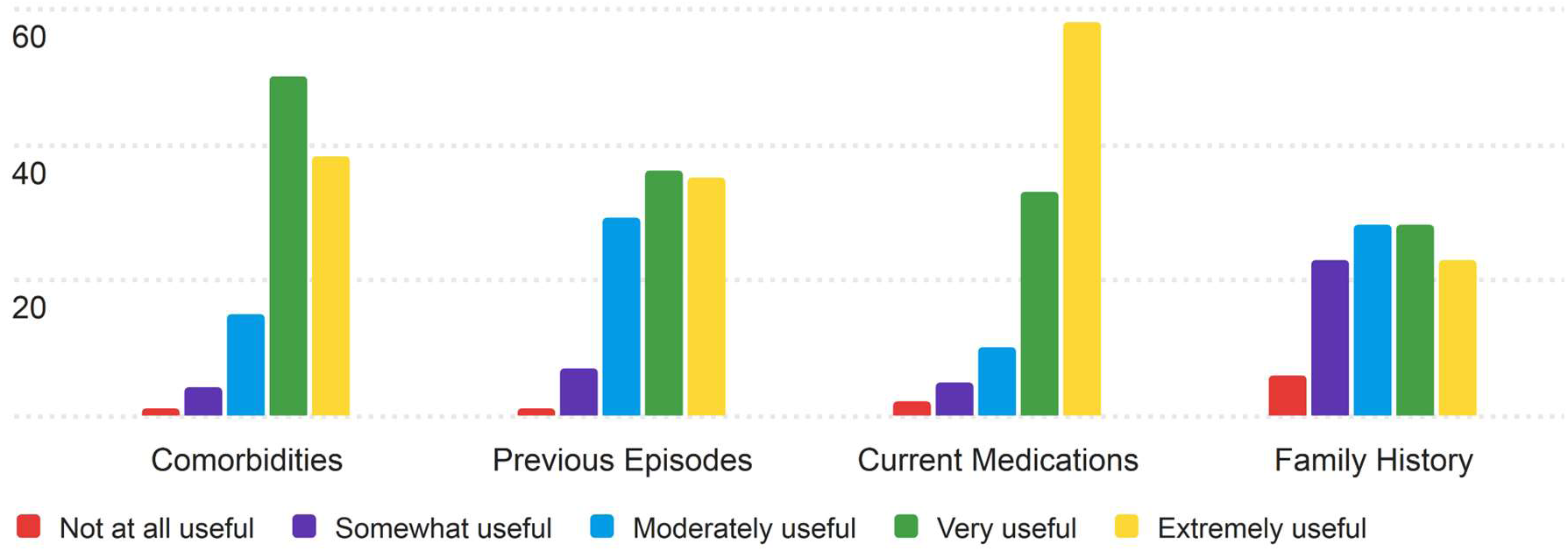
Response Distribution for the Usefulness of the Background section.

As mentioned above, the distribution of answers for Previous Episodes and Family History both see more grouping around 4 on the Likert scale (very useful) and see more nurses rating these sections at a 3 (moderately useful) and less rating them 5 (extremely useful).

#### 3.1.6 Actions

The sixth section of the PRT that nurses were asked to rate for its usefulness was the Actions section. The results of the nurses’ ratings are displayed in Table 15.

**Table 15.** Nurses’ Ratings of the Actions section.

| Actions | Mode | Median | Average | Minimum | Maximum | Count |
| --- | --- | --- | --- | --- | --- | --- |
| Actions Taken: Actions and interventions that have already been completed | 4 | 4 | 4.03 | 1 | 5 | 106 |
| Actions required/ongoing: Explain any treatments or procedures currently in progress or that need to be completed still | 5 | 4 | 4.08 | 1 | 5 | 106 |
| Rationale: Justification for previous and current/future actions | 4 & 5 | 4 | 3.71 | 1 | 5 | 105 |
| Signs to elevate: Symptoms which indicate need to alert physician/nurse practitioner | 5 | 4 | 4.17 | 1 | 5 | 105 |

Generally, this sample of nurses found the Actions section to be useful, with most categories achieving an average score above 4 (Very Useful) on the 5-point Likert scale. Signs to elevate had the highest average score (4.17), with a mode of 5 (Extremely Useful), suggesting that nurses found this category particularly valuable. Actions required/ongoing also received a strong average score of 4.08.

The Rationale category had the lowest average score (3.71), indicating a more mixed perception of its usefulness, with a broader distribution of responses. Most categories, including Actions Taken, had a median of 4, reflecting that many nurses found these sections "Very Useful" in their daily work. The distributions of responses in the Actions section are displayed in Figure 6.

**Figure 6.**
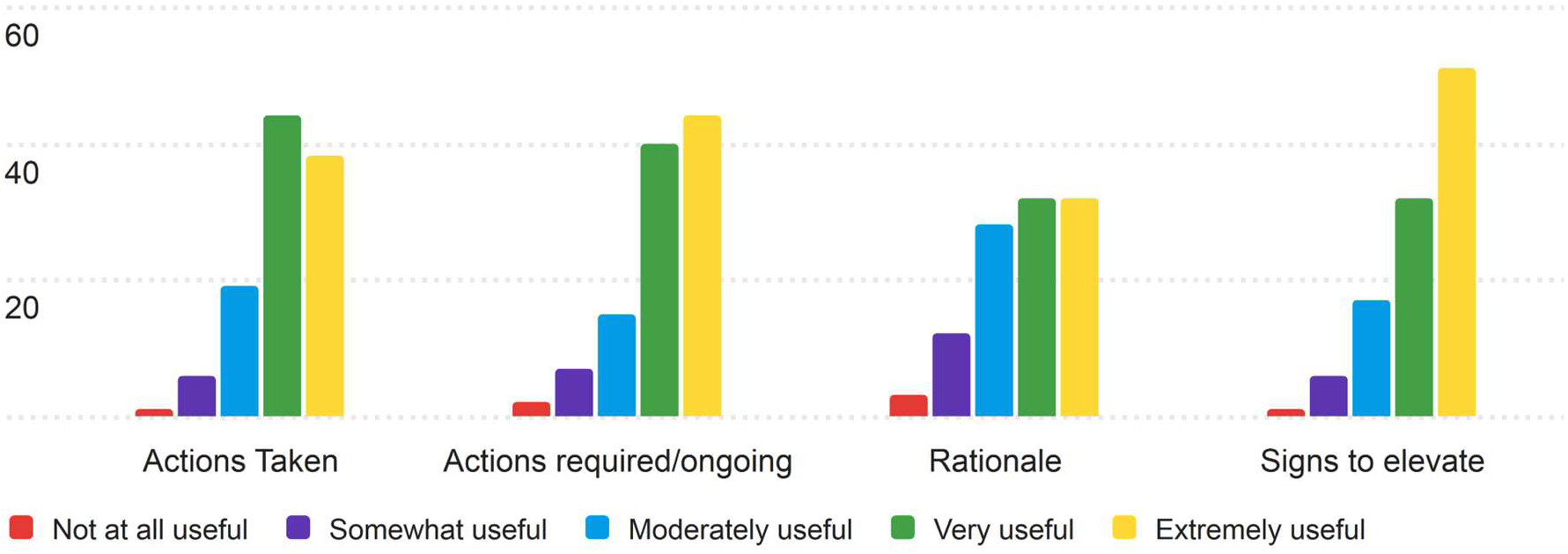
Response Distribution for the Usefulness of the Actions Section.

The distribution of ratings shows that "Very Useful" and "Extremely Useful" were the most common responses across all categories, with a few lower ratings in the "Somewhat Useful" range, particularly for the Rationale category.

#### 3.1.7 Timing

The seventh section of the PRT that nurses were asked to rate for its usefulness was the Timing section. The results of the nurses’ ratings are displayed in Table 16.

**Table 16.** Nurses’ Ratings of the Timing section.

| Timing | Mode | Median | Average | Minimum | Maximum | Count |
| --- | --- | --- | --- | --- | --- | --- |
| Levels of Urgency/ Prioritization: Order of actions of care in decreasing priority | 5 | 4 | 3.88 | 1 | 5 | 106 |
| Explicit Timing: If specific timestamps for certain actions (administer medication, remove ventilator) | 5 | 4 | 3.94 | 1 | 5 | 106 |
| Coordination: Mention any coordination needed with other departments or services | 4 | 4 | 3.72 | 1 | 5 | 106 |

Nurses found the Timing section to be useful, with average scores between 3.72 and 3.94. Explicit Timing received the highest average score of 3.94, indicating its relevance to nurses in their day-to-day workflows. Levels of Urgency/Prioritization also performed well, with an average score of 3.88, while Coordination had the lowest average score (3.72), reflecting a slightly more mixed perception of its usefulness. The full distribution of responses is displayed in Figure 7.

**Figure 7.**
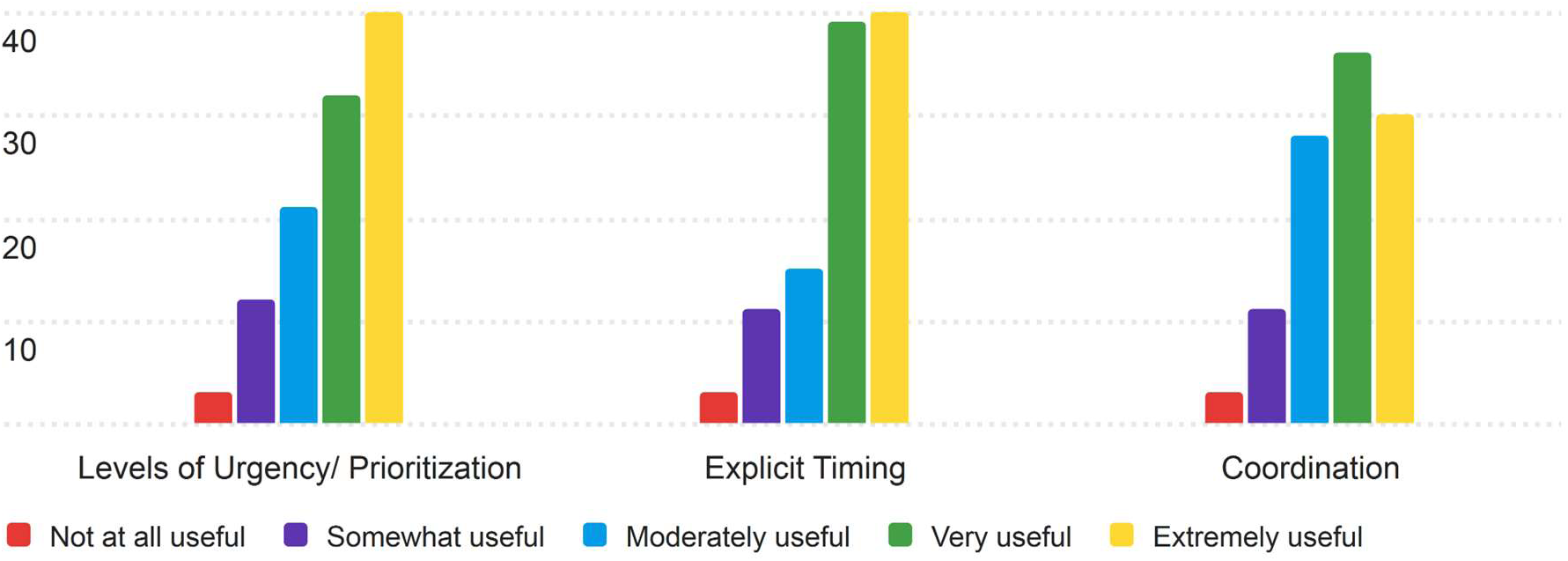
Response Distribution for the Usefulness of the Timing section.

Most ratings for the Timing categories clustered around 4 (Very Useful), with the most common mode being 5 (Extremely Useful) for Levels of Urgency and Explicit Timing. Coordination had a wider range of ratings, with more responses in the "Moderately Useful" and "Very Useful" ranges.

#### 3.1.8 Ownership

The eighth section of the PRT that nurses were asked to rate for its usefulness was the Ownership section. The results of the nurses’ ratings are displayed in Table 17.

**Table 17.** Nurses’ ratings of the Ownership section.

| Ownership | Mode | Median | Average | Minimum | Maximum | Count |
| --- | --- | --- | --- | --- | --- | --- |
| Responsibility: What person/team in charge of care | 5 | 4 | 4.19 | 1 | 5 | 105 |
| Patient/Family: Who is making medical decisions for patient | 5 | 4 | 3.95 | 1 | 5 | 105 |

For the ownership section, nurses found the components to be useful, with both categories scoring averages of 3.95 and 4.19 on the 5-point Likert scale respectively. Responsibility received the highest average score of 4.19, indicating that nurses considered this information potentially useful. Patient/Family, while still rated as useful, had a slightly lower average of 3.95. Figure 8 contains the distribution of responses for the ownership section.

**Figure 8.**
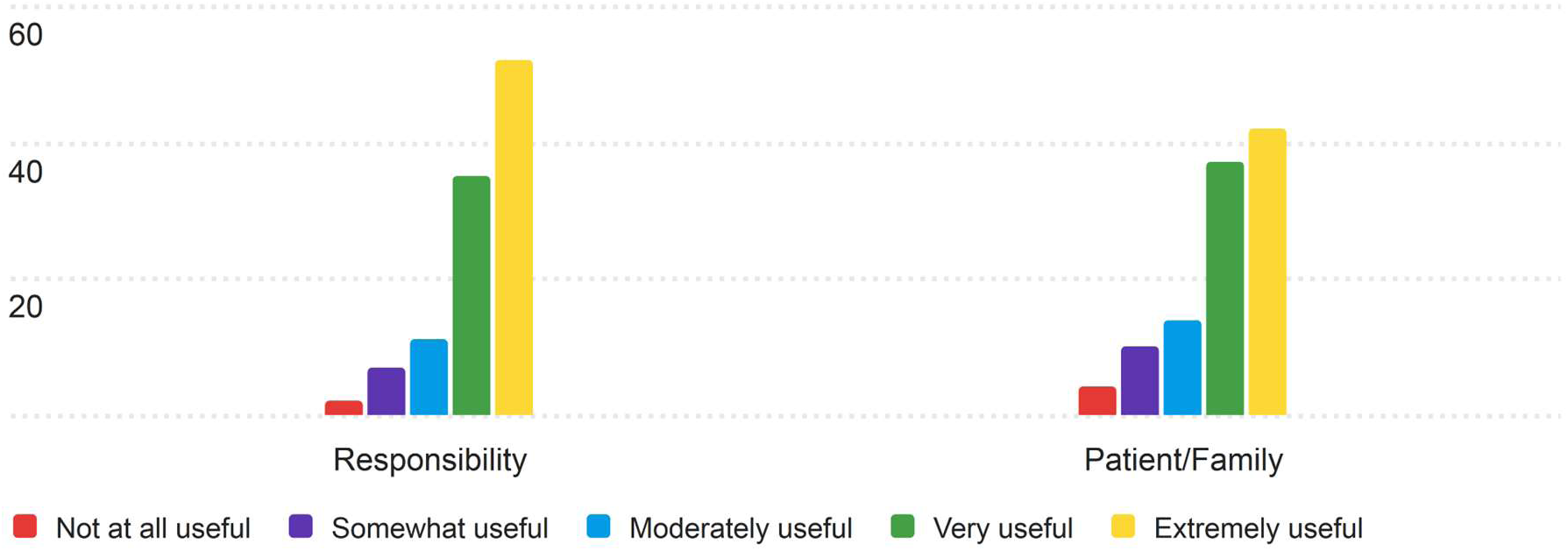
Response Distribution for the Usefulness of the Ownership section.

In both categories, the most common (mode) rating was 5 (Extremely Useful), and the median score for both categories was 4 (Very Useful). The distribution of ratings shows a concentration of responses in the "Very Useful" and "Extremely Useful" ranges for both categories.

#### 3.1.9 Next

The ninth section of the PRT that nurses were asked to rate for its usefulness was the Next section. The results of the nurses’ ratings are displayed in Table 18.

**Table 18.** Nurses’ ratings of the Next section.

| Next | Mode | Median | Average | Minimum | Maximum | Count |
| --- | --- | --- | --- | --- | --- | --- |
| Anticipated Changes: Any possible/likely changes in status for patients because of interventions/care | 4 | 4 | 3.8 | 1 | 5 | 106 |
| Plan of action: If current conditions persist, plan of care for patient | 5 | 4 | 3.98 | 1 | 5 | 106 |
| Contingency Plans: If current conditions change in specific ways, adaptations to plan or other plans of care for the patient | 4 | 4 | 3.95 | 1 | 5 | 106 |

Overall, nurses found the Next section to be useful, with average scores between 3.8 and 3.98. Plan of Action received the highest average score (3.98) and a mode of 5 (Extremely Useful), reflecting its perceived importance among nurses. Contingency Plans followed closely with an average score of 3.95, while Anticipated Changes had the lowest average (3.80), although it was still considered useful by most respondents. The distribution of responses are displayed in Figure 9.

**Figure 9.**
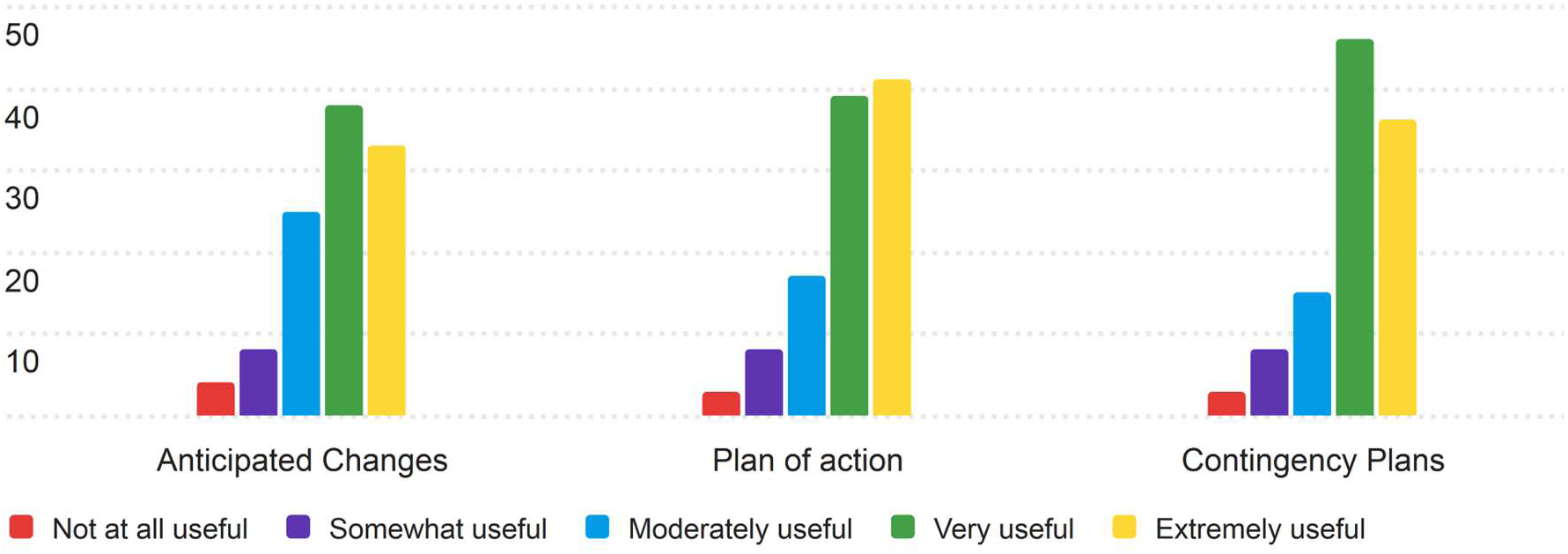
Response Distribution for the Usefulness of the Next section.

Most ratings across all categories were concentrated around 4 (Very Useful), with fewer responses in the "Somewhat Useful" and "Not at all Useful" ranges. This suggests that nurses generally thought the components of the Next would be useful in their daily workflows.

### 3.2 AI Sentiment

Besides rating the PRT, nurses were also asked three questions about their perceptions of AI in their daily work. Nurses were asked to evaluate their Comfort, Trust, and expected utility in their daily work. They answered based on 5-point Likert scales, with 1 being the least comfortable / trusting / valuable, and 5 being the most. Nurses’ opinions on these matters are shown in table 19.

**Table 19.** Nurses’ ratings for Perceptions of AI.

| Perceptions of AI | Mode | Median | Average | Minimum | Maximum | Count |
| --- | --- | --- | --- | --- | --- | --- |
| Comfort using AI in your work (1-5) | 2 | 2 | 2.38 | 1 | 4 | 105 |
| Level of Trust (1-5) | 2 | 2 | 2.5 | 1 | 5 | 105 |
| Expected Utility (1-5) | 3 | 3 | 3 | 1 | 5 | 106 |

Nurses exhibited relatively low levels of trust and comfort when it comes to AI in their workflows, with no nurses indicating that they were Extremely comfortable (Likert score of 5) with using AI generated reports in their day-to-day work. For both Comfort and trust, median and mode values of 2 indicate low overall opinions in these categories, with expected utility faring slightly better, with mean, median, and mode all scoring 3, or moderate utility, on the survey. Below are the distributions of score for each of the above questions. The distribution of responses for comfort is shown in Figure 10.

**Figure 10.**
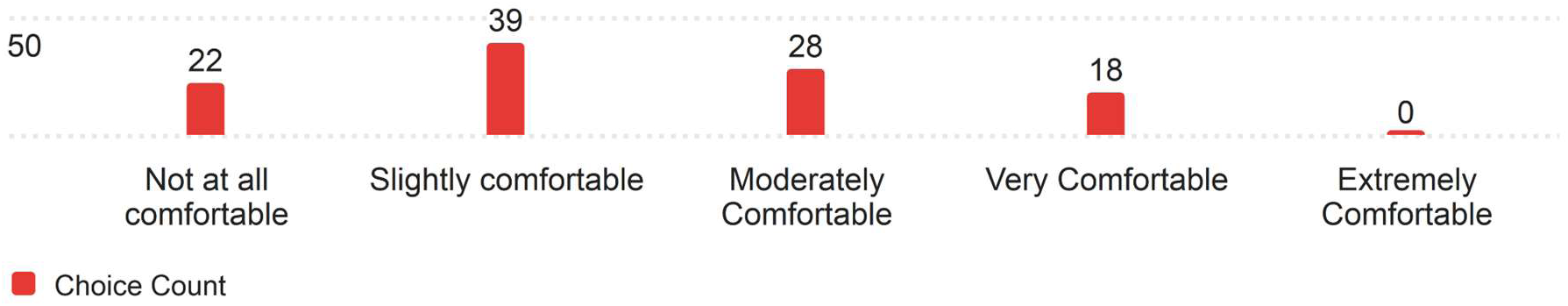
Response Distribution for Comfort using AI (n= 107)

Most nurses surveyed felt slightly comfortable using AI generated patient reports in their work, with a comparable number of nurses feeling both Not at all comfortable (1 on the Likert scale) and moderately comfortable (3 on the Likert scale) with using AI patient reports. A minority of nurses felt Very comfortable (4 on the Likert scale) and none felt extremely comfortable. The distribution of responses for the level of trust in AI are displayed in Figure 11.

**Figure 11.**
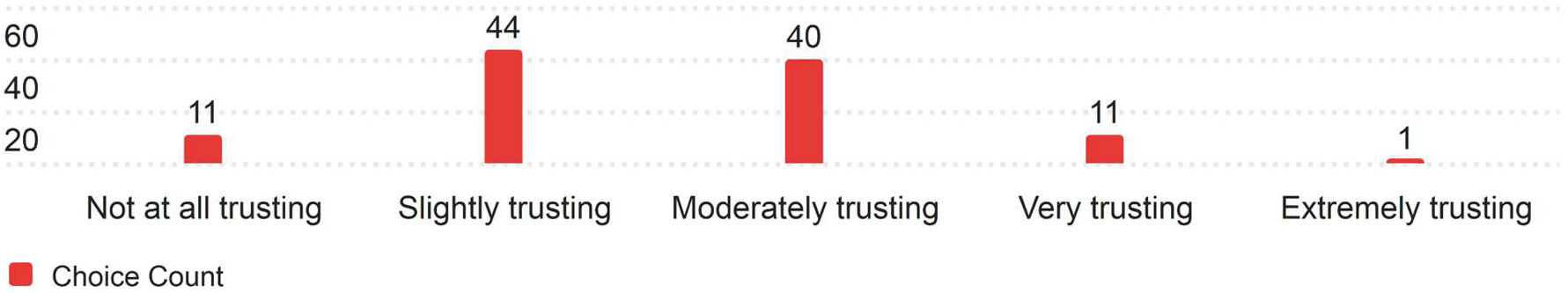
Response Distribution for Level of Trust in AI (n=107)

Most answers in regard to trust with AI generated patient templates centered around 2 and 3 on the Likert scale (slightly and moderately trusting of AI patient reports), with an equal number of nurses feeling both not at all trusting and very trusting (1 and 4 on the Likert scale) respectively. Only one nurse was extremely trusting of AI patient reports for their day-to-day work. Figure 12 displays Nurses’ expected utility from AI in their workplace.

**Figure 12.**
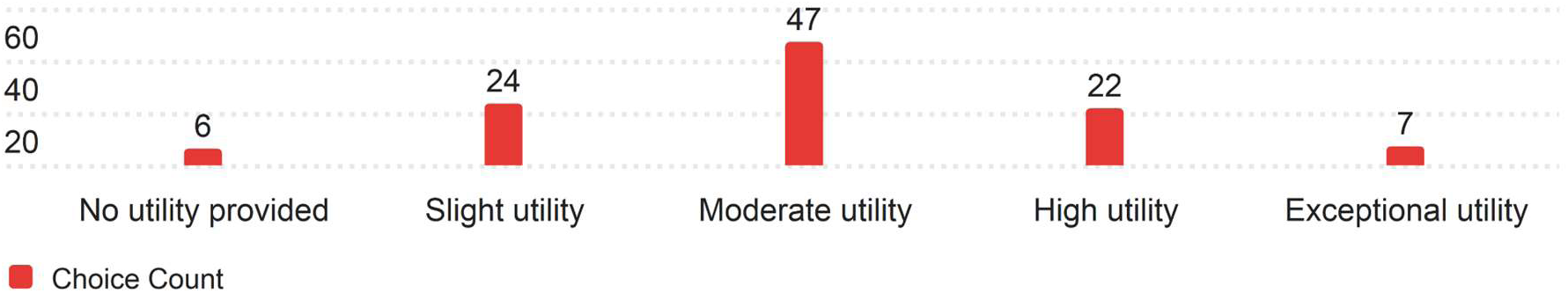
Response Distribution for Expected utility from AI (n= 108)

When it comes to nurses’ expectations of utility from AI patient generated reports, most nurses believe that they can expect moderate utility (3 on the Likert scale) from such reports. There is a comparable number of nurses who expect slight or high utility (2 and 4 on the Likert scales) and a relatively small number of nurses who expect either no utility or exceptional utility (1 and 5 on the Likert scale, respectively).

### 3.3 Population

Nurses were asked to give some basic demographic information, pertaining to their years of experience, age, and primary nursing unit of work. For Years of experience, nurses chose from 3 ranges: 0-10 Years, 11-20 Years, and 21-30 Years of experience. For Age, nurses chose from 3 ranges: Young Adult (18-35), Adult (36-59), and Older Adult (60+). For the question about a Nurse’s unit, they were asked to choose from the list of nursing departments listed on the University of Iowa Health Care’s Website. The results are displayed in Table 20 for Department, Table 21 for Years of Experience, and Table 22 for Age.

**Table 20.**
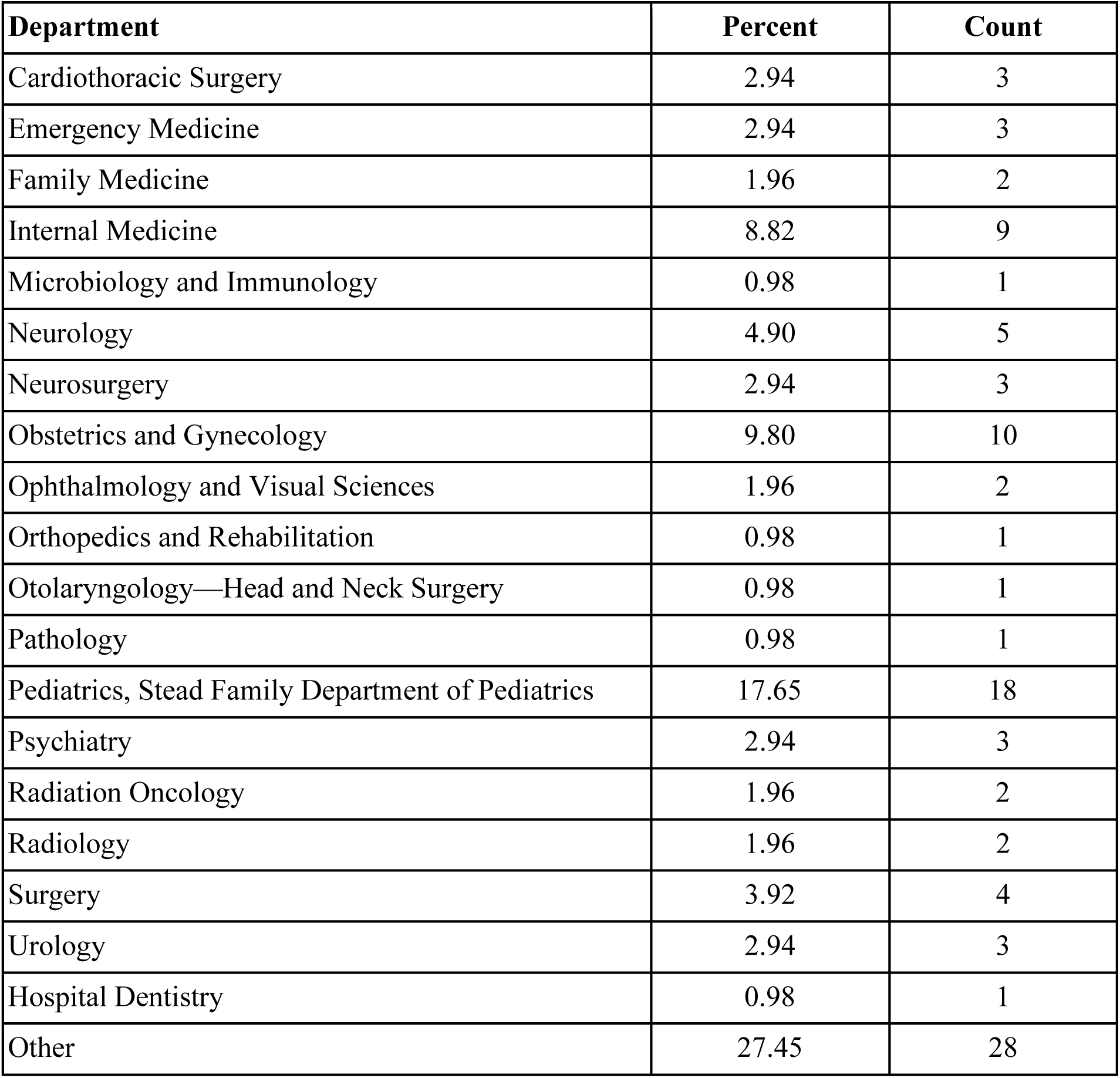
Composition by Unit of sampled Nurse Population.

| Department | Percent | Count |
| --- | --- | --- |
| Cardiothoracic Surgery | 2.94 | 3 |
| Emergency Medicine | 2.94 | 3 |
| Family Medicine | 1.96 | 2 |
| Internal Medicine | 8.82 | 9 |
| Microbiology and Immunology | 0.98 | 1 |
| Neurology | 4.90 | 5 |
| Neurosurgery | 2.94 | 3 |
| Obstetrics and Gynecology | 9.80 | 10 |
| Ophthalmology and Visual Sciences | 1.96 | 2 |
| Orthopedics and Rehabilitation | 0.98 | 1 |
| Otolaryngology—Head and Neck Surgery | 0.98 | 1 |
| Pathology | 0.98 | 1 |
| Pediatrics, Stead Family Department of Pediatrics | 17.65 | 18 |
| Psychiatry | 2.94 | 3 |
| Radiation Oncology | 1.96 | 2 |
| Radiology | 1.96 | 2 |
| Surgery | 3.92 | 4 |
| Urology | 2.94 | 3 |
| Hospital Dentistry | 0.98 | 1 |
| Other | 27.45 | 28 |

**Table 21.**
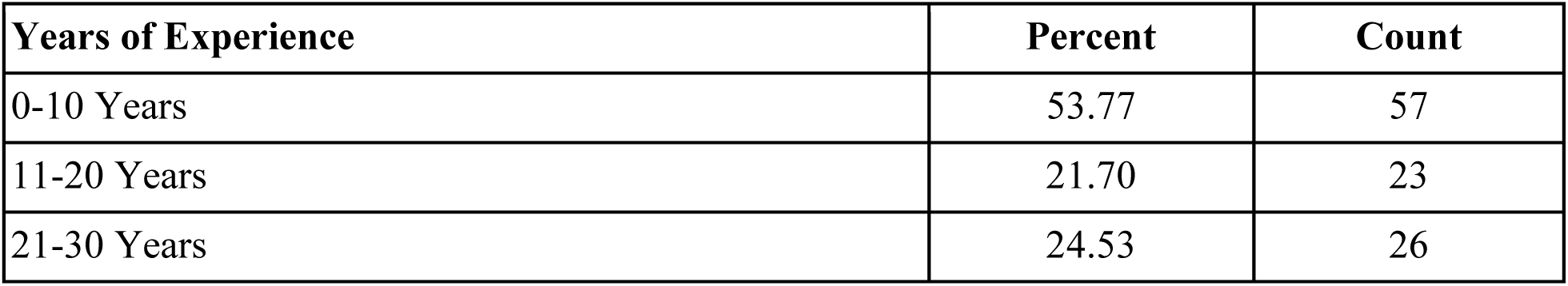
Composition by years of experience of sampled nurse population.

| Years of Experience | Percent | Count |
| --- | --- | --- |
| 0-10 Years | 53.77 | 57 |
| 11-20 Years | 21.70 | 23 |
| 21-30 Years | 24.53 | 26 |

**Table 22.** Composition by age of sampled Nurse population.

| Age | Percent | Count |
| --- | --- | --- |
| Young Adult (18-35) | 44.44 | 48 |
| Adult (36-59) | 46.30 | 50 |
| Older Adult (60+) | 9.26 | 10 |

The most represented types of nurses for this study were Pediatrics at 18% (n=18), Obstetrics and Gynecology at 10% (n=10), and Internal Medicine at 9% of survey respondents (n=9). There was a notable number of nurses who consider themselves to be in the other category at 27% (n=28)

Over half of the nurses who responded are within their first 10 years of work as a nurse (∼54%, n=57), with a roughly even spread between 11-20 and 21-30 years of experience (∼22%, n=23 & ∼24%, n=26 respectively).

The population who was surveyed for this study had a roughly even amount of Young Adult and Adult nurses (n=48 and 50 respectively), and a relatively small population of Older Adult nurses with 10 nurses who match that category responding to the survey. While small, the input of older nurses with the most experience is valuable to the study.

## 4. Discussion

This analysis discusses the perceived usefulness of the PRT from the perspective of practicing nurses. Overall, nurses expected most components of the PRT to be useful in their daily workflow. Further, nurses also gave their opinions related to the level of trust, comfort, and expected utility of AI in their daily workflow. Nurses are neither trusting nor comfortable currently using AI, but they do expect a high amount of utility to be provided by future AI based tools and systems.

### 4.1 Patient Report Template

#### Patient Profile

The Patient Profile section of the PRT received consistently high ratings, with all components scoring an average above 4, indicating perceived usefulness during patient handoffs. Patient’s Name and Code Status were rated highest, with averages of 4.42 and 4.38, reflecting the importance of quick access to key identification and critical status details. While Sex received the lowest average rating (4.03), it still fell within the "Very Useful" range, underscoring its relevance. The overall high scores suggest a consensus among these nurses that the Patient Profile enhances access to essential patient information. Further research is needed to assess how this perceived usefulness impacts workflow and patient care outcomes.

#### Review of Systems

The Review of Systems section received varied ratings, with Chief Complaint rated highest (4.40) and Eyes lowest (3.73). Nurses generally found this section useful, particularly components like Chief Complaint, Cardiovascular (4.23), and Respiratory (4.24), which align with their clinical importance. Lower scores for Eyes, Integumentary, and Musculoskeletal (all below 4.0) suggest these areas may be less central to nursing workflows or vary in relevance based on patient population or clinical context. A mode of 5 (Extremely Useful) across all sections indicates broad utility, though some variability in responses highlights the need for adaptability. Refining the template to allow customization of less-used sections could enhance its relevance for diverse patient scenarios and nursing roles.

#### Situation

The Situation section received mixed ratings, with averages ranging from 3.25 to 3.96. Recent Changes and Plan of Care scored highest (median 5), reflecting their relevance to nursing workflows by providing actionable, real-time patient information. Conversely, Level of Uncertainty had the lowest average (3.25), with some nurses finding it less applicable, as quantifying uncertainty may not align with their emphasis on direct and clear communication during handoffs. The variability in responses, including some low ratings, suggests that while the section is broadly useful, components like Level of Uncertainty may require refinement or reconsideration to better fit practical nursing needs.

#### Safety

The Safety section received high ratings, with all components—Lab Values, Allergies, and Alerts—scoring averages above 4 (Very Useful). Allergies scored the highest (4.36), with a strong consensus on its critical importance for preventing adverse reactions. Lab Values (4.21) and Alerts (4.22) also performed well, with most responses in the "Very Useful" or "Extremely Useful" range. These results highlight the importance of accessible, organized safety information during handoffs, aligning with nurses’ priorities for tools that support patient safety and error prevention.

#### Background

The Background section received mixed ratings, with averages ranging from 3.36 (Family History) to 4.29 (Current Medications). Current Medications was rated the highest, reflecting their critical importance in nursing workflows. Comorbidities (4.11) and Previous Episodes (3.91) were also valued for providing key medical history. Family History scored the lowest (3.36), indicating moderate usefulness with variability based on context. These results suggest that while the Background section is broadly useful, its relevance could be enhanced by tailoring components to specific patient populations or workflows.

#### Actions

The Actions section of the PRT was generally well received, with most categories scoring above 4 (Very Useful). "Signs to Elevate" ranked highest (4.17), reflecting its importance in supporting timely care escalation. "Actions Required/Ongoing" also scored well (4.08), emphasizing the value of tracking incomplete treatments. In contrast, "Rationale" received the lowest average (3.71), indicating mixed feedback, as some nurses may prioritize immediate tasks over detailed explanations. While "Very Useful" and "Extremely Useful" were the most frequent responses, variability in the "Rationale" category highlights differences in its perceived importance across nursing contexts.

#### Timing

The Timing section of the PRT received positive feedback, with most components rated as useful. "Explicit Timing" scored the highest (3.94), reflecting its relevance for time-sensitive tasks like medication administration. "Levels of Urgency/Prioritization" also received strong ratings (3.88), highlighting the value of clear task prioritization during shift changes. "Coordination," which relates to interdepartmental communication, received the lowest average (3.72), indicating more mixed perceptions, possibly reflecting variability in nursing roles and work environments. The Timing section was valued for helping prioritize tasks and ensure timely actions.

#### Ownership

The Ownership section of the PRT was well-received, with nurses rating "Responsibility" (4.19) higher than "Patient/Family" (3.95) on a 5-point Likert scale. Both categories had a mode of 5 ("Extremely Useful") and a median of 4 ("Very Useful"), indicating general agreement on their value. However, "Patient/Family" showed slightly more variability in responses. Overall, the section was seen by this group as beneficial for clarifying care responsibility and medical decision-making, supporting improved communication during patient handoffs.

#### Next

The "Next" section was rated as useful, with average scores ranging from 3.80 to 3.98. "Plan of Action" received the highest average (3.98) and a mode of 5, highlighting its importance in guiding care continuity when conditions persist. "Contingency Plans" scored 3.95, emphasizing the value of having structured alternatives for unexpected patient changes. "Anticipated Changes" had the lowest average (3.80) and a mode of 4, indicating its usefulness but less central role compared to planning and contingency. Ratings were concentrated around 4 (Very Useful), reflecting general agreement on the section’s relevance, with the reduced perceived importance of anticipating future changes.

### 4.2 AI Sentiment

The survey also explored nurses’ sentiments toward integrating AI into their workflows, focusing on Comfort, Trust, and Expected Utility. Overall, responses revealed a general skepticism toward AI technologies. Comfort with AI was low, with a mode and median of 2 (slightly comfortable). A significant portion of nurses (22) reported no comfort at all, while 18 expressed being very comfortable, and none felt extremely comfortable. Trust levels followed a similar pattern, with 44 nurses slightly trusting AI (mode = 2) and only 11 expressing very high trust. One respondent rated trust as extremely high. This hesitation may reflect concerns about reliability, transparency, and the implications of AI-driven decision-making in clinical settings. In contrast, Expected Utility showed more balanced views. Most nurses (47) anticipated moderate utility, with smaller groups expecting high (22) or slight utility (24). Only 7 saw exceptional utility, while 6 saw none. These mixed responses indicate cautious recognition of AI’s potential, tempered by the need for further evidence of its effectiveness in improving workflows and patient outcomes. Addressing barriers to AI adoption will require greater transparency, demonstrated reliability, and seamless integration into workflows. Ensuring that AI tools support rather than burden nurses is essential for fostering trust and comfort.

### 4.3 Population Information

The study respondents represented a diverse range of departments, with Pediatrics comprising the largest group (18%), followed by Obstetrics and Gynecology (10%) and Internal Medicine (9%). Notably, 27% identified working in "other" departments, indicating a significant presence of nurses from niche or specialized fields. This broad departmental representation supports the generalizability of the findings while suggesting that perceptions of the PRT’s usefulness may vary by clinical setting. Customizing the PRT to meet the unique demands of specific specialties may enhance its relevance and utility. Over half of the respondents (54%) had 0-10 years of nursing experience, indicating a majority (early in their careers) who may be more open to adopting structured tools like the PRT.

The remaining participants were evenly split between 11-20 years (22%) and 21-30 years (24%) of experience, bringing insights from more seasoned professionals who may critically evaluate how new tools align with established workflows. The respondents’ age distribution was nearly equal between Young Adults (44%) and Adults (46%), with Older Adults representing a smaller share (9%). Younger nurses, likely more accustomed to digital tools, may be more receptive to innovations like the PRT, while older, experienced nurses provide valuable perspectives on potential barriers and workflow integration. The diversity in department, experience, and age highlights the need for a customizable and adaptable PRT to address the distinct requirements of various clinical environments and nursing roles.

### 4.4 Limitations

This study has several limitations that may have influenced the results. Selection bias is a key concern, as the survey was limited to nurses within the University of Iowa Health Care system, potentially reducing the generalizability of findings to institutions with different workflows or structures. Response bias, particularly social desirability bias, may also have affected results, with nurses potentially providing responses they believed aligned with researchers’ expectations regarding workflow and patient care improvements. To address these biases, the PRT was developed using established nurse handoff guidelines, such as SBAR and I PASS the BATON, ensuring alignment with recognized best practices. Additionally, the survey sought detailed feedback on individual PRT components rather than relying on generalized ratings, encouraging critical evaluation of the tool’s features. Another limitation is the survey’s reliance on nurses’ perceptions of expected usefulness rather than hands-on testing of the PRT. Without real-world interaction, responses may have been overly speculative. Future research should integrate the PRT into clinical practice to gather feedback based on actual use, offering a more accurate evaluation of its impact on nursing workflows.

## 5. Conclusion

The feedback from nurses highlights the potential of the PRT to improve nursing workflows, particularly during patient handoffs. Highly rated components, such as the Patient Profile, Actions, and Safety sections, indicate that nurses view the PRT as a potentially valuable tool for streamlining information transfer and reducing charting time. Its alignment with established handoff frameworks like SBAR and I PASS the BATON further supports its relevance in clinical workflows. The PRT also addresses key inefficiencies documented in the literature, such as EHR-related cognitive load and charting burdens. By providing a structured format that enhances clarity and accuracy, the PRT may reduce handoff errors and improve patient safety outcomes— critical priorities in nursing practice.

Future research should focus on testing the PRT in clinical settings to validate these findings through hands-on use. Specialized nursing units, such as elderly or memory care, could provide additional insights into the template’s adaptability. Emphasis should also be placed on usability testing and validating the accuracy of AI-generated reports to ensure seamless integration into existing workflows. Addressing these areas will enhance the PRT’s impact on improving efficiency and patient care across diverse clinical environments.

## Data Availability

All data produced in the present work are contained in the manuscript

## Acknowledgements

The authors would like to thank Dadriane Fice for her insights on this study.

## Declaration of Generative AI and AI-Assisted Technologies

During the preparation of this work, the authors used ChatGPT to improve the flow of the text, correct any potential grammatical errors, and improve the writing. After using this tool, the authors reviewed and edited the content as needed and took full responsibility for the content of the publication.

## Credit Author Statement

**Gabriel Vald**: Conceptualization, Methodology, Software, Validation, Formal analysis, Investigation, Data Curation, Writing - Original Draft, and Visualization. **Yusuf Sermet**: Conceptualization, Methodology, Writing - Review & Editing, Investigation, Validation, Supervision. **Nai-Ching Chi**: Validation, Writing - Review & Editing, **Ibrahim Demir**: Project administration, Writing - Review & Editing, Funding acquisition, and Resources.

## References

American College of Cardiology. (n.d.). Review of Systems. https://www.acc.org/Tools-and-Practice-Support/Practice-Solutions/Coding-and-Reimbursement/Documentation/Evaluation-and-Management/Review-of-Systems

Ash, J. S., Sittig, D. F., Campbell, E. M., Guappone, K. P., & Dykstra, R. H. (2007). Some unintended consequences of clinical decision support systems. AMIA … Annual Symposium proceedings. AMIA Symposium, 2007, 26–30.

Devaraj, S., Sharma, S. K., Fausto, D. J., Viernes, S., & Kharrazi, H. (2014). Barriers and facilitators to Clinical Decision Support Systems Adoption: A systematic review. Journal of Business Administration Research, 3(2). 10.5430/jbar.v3n2p36

Dowding, D., Mitchell, N., Randell, R., Foster, R., Lattimer, V., & Thompson, C. (2009). Nurses’ use of computerised clinical decision support systems: A case site analysis. Journal of Clinical Nursing, 18(8), 1159–1167. 10.1111/j.1365-2702.2008.02607.x

Dunn Lopez, K., Gephart, S. M., Raszewski, R., Sousa, V., Shehorn, L. E., & Abraham, J. (2016). Integrative Review of Clinical Decision Support for registered nurses in Acute Care Settings. Journal of the American Medical Informatics Association, 24(2), 441–450. 10.1093/jamia/ocw084

Effken, J., Loeb, R., Kang, Y., & Lin, Z. (2008). Clinical information displays to improve ICU outcomes. International Journal of Medical Informatics, 77(11), 765–777. 10.1016/j.ijmedinf.2008.05.004

Gardner, R. L., Cooper, E., Haskell, J., Harris, D. A., Poplau, S., Kroth, P. J., & Linzer, M. (2018). Physician stress and burnout: The impact of health information technology. Journal of the American Medical Informatics Association, 26(2), 106–114. 10.1093/jamia/ocy145

Keenan, G., Yakel, E., Dunn Lopez, K., Tschannen, D., & Ford, Y. B. (2013). Challenges to nurses’ efforts of retrieving, documenting, and Communicating Patient Care Information. Journal of the American Medical Informatics Association, 20(2), 245–251. 10.1136/amiajnl-2012-000894

Kossman, S. P., & Scheidenhelm, S. L. (2008). Nurses’ perceptions of the impact of electronic health records on work and patient outcomes. *CIN: Computers, Informatics*, Nursing, 26(2), 69–77. 10.1097/01.ncn.0000304775.40531.67

Liang, Y.-W., Chen, W.-Y., Lee, J.-L., & Huang, L.-C. (2012). Nurse staffing, direct nursing care hours and patient mortality in Taiwan: The Longitudinal Analysis of hospital nurse staffing and patient outcome study. BMC Health Services Research, 12(1). 10.1186/1472-6963-12-44

Park, H.-A., Murray, P. J., & Delaney, C. W. (2006). Consumer-centered computer-supported care for Healthy People: Proceedings of NI2006, the 9th International Congress on Nursing Informatics. IOS Press.

Pursnani, V., Sermet, Y., Kurt, M., & Demir, I. (2023). Performance of ChatGPT on the US fundamentals of engineering exam: Comprehensive assessment of proficiency and potential implications for professional environmental engineering practice. Computers and Education: Artificial Intelligence, 5, 100183.

Riesenberg, L. A., Leisch, J., & Cunningham, J. M. (2010). Nursing handoffs: A systematic review of the literature. *AJN*, American Journal of Nursing, 110(4), 24–34. 10.1097/01.naj.0000370154.79857.09

Sajja, R., Sermet, Y., Cwiertny, D., & Demir, I. (2023a). Platform-independent and curriculum-oriented intelligent assistant for higher education. International Journal of Educational Technology in Higher Education, 20(1), 42.

Sajja, R., Sermet, Y., Cwiertny, D., & Demir, I. (2023b). Integrating AI and Learning Analytics for Data-Driven Pedagogical Decisions and Personalized Interventions in Education. arXiv preprint arXiv:2312.09548.

Samuel, D. J., Sermet, M. Y., Mount, J., Vald, G., Cwiertny, D., & Demir, I. (2024). Application of Large Language Models in Developing Conversational Agents for Water Quality Education, Communication and Operations. EarthArxiv, 7056. 10.31223/X5XT4K

Schachner, M. B., Recondo, F. J., Sommer, J. A., González, Z. A., García, G. M., Luna, D. R., & Benítez, S. E. (2015). Pre-implementation study of a nursing e-chart: How nurses use their time. In MEDINFO 2015: eHealth-enabled Health (pp. 255-258). IOS Press.

Sermet, Y., & Demir, I. (2021). A semantic web framework for automated smart assistants: A case study for public health. Big Data and Cognitive Computing, 5(4), 57.

Shahid, S., & Thomas, S. (2018). Situation, background, assessment, recommendation (SBAR) communication tool for handoff in Health Care – A Narrative Review. Safety in Health, 4(1). 10.1186/s40886-018-0073-1

Sutton, R. T., Pincock, D., Baumgart, D. C., Sadowski, D. C., Fedorak, R. N., & Kroeker, K. I. (2020). An overview of clinical decision support systems: Benefits, risks, and strategies for Success. Npj Digital Medicine, 3(1). 10.1038/s41746-020-0221-y

Tang, L., Sun, Z., Idnay, B., Nestor, J. G., Soroush, A., Elias, P. A., Xu, Z., Ding, Y., Durrett, G., Rousseau, J. F., Weng, C., & Peng, Y. (2023). Evaluating large language models on medical evidence summarization. Npj Digital Medicine, 6(1). 10.1038/s41746-023-00896-7

Titles - H.R.1 - 111th Congress (2009-2010): American Recovery and Reinvestment Act of 2009. (2009, February 17). https://www.congress.gov/bill/111th-congress/house-bill/1/titles

Text - H.R.2 - 114th Congress (2015-2016): Medicare Access and CHIP Reauthorization Act of 2015. (2015, April 16). https://www.congress.gov/bill/114th-congress/house-bill/2/text

UIHC. (n.d.). Basic facts about medical center on the university campus. University of Iowa Health Care. https://uihc.org/basic-facts-about-medical-center-university-campus

Van Veen, D., Van Uden, C., Blankemeier, L., Delbrouck, J.-B., Aali, A., Bluethgen, C., Pareek, A., Polacin, M., Reis, E. P., Seehofnerová, A., Rohatgi, N., Hosamani, P., Collins, W., Ahuja, N., Langlotz, C. P., Hom, J., Gatidis, S., Pauly, J., & Chaudhari, A. S. (2024). Adapted large language models can outperform medical experts in clinical text summarization. Nature Medicine, 30(4), 1134–1142. 10.1038/s41591-024-02855-5

Yee, T., Needleman, J., Pearson, M., Parkerton, P., Parkerton, M., & Wolstein, J. (2012). The influence of integrated electronic medical records and computerized nursing notes on nurses’ time spent in documentation. CIN: Computers, Informatics, Nursing, 1. 10.1097/nxn.0b013e31824af835

Yen, P. Y., Kellye, M., Lopetegui, M., Saha, A., Loversidge, J., Chipps, E. M., Gallagher-Ford, L., & Buck, J. (2018). Nurses’ Time Allocation and Multitasking of Nursing Activities: A Time Motion Study. AMIA … Annual Symposium proceedings. AMIA Symposium, 2018, 1137–1146.

Zhang, M., Zhu, L., Lin, S.Y., Herr, K., Chi, C.L., Demir, I., Dunn Lopez, K. and Chi, N.C., (2023). Using artificial intelligence to improve pain assessment and pain management: a scoping review. Journal of the American Medical Informatics Association, 30(3), pp.570–587.

Zhang, T., Ladhak, F., Durmus, E., Liang, P., McKeown, K., & Hashimoto, T. B. (2024). Benchmarking large language models for news summarization. Transactions of the Association for Computational Linguistics, 12, 39–57. 10.1162/tacl_a_00632

